# How early can an upcoming critical transition be detected?

**DOI:** 10.1101/2022.05.27.22275693

**Authors:** Emma Southall, Michael J Tildesley, Louise Dyson

## Abstract

Numerous studies have suggested the use of early warning signals (EWSs) of critical transitions to overcome challenges of identifying tipping points in complex natural systems. However, the real-time application of EWSs has often been overlooked; many studies show the presence of EWSs but do not detect when the trend becomes significant. Knowing if the signal can be detected *early* enough is of critical importance for the applicability of EWSs. Detection methods which present this analysis are sparse and are often developed anew for each individual study. Here, we provide a summary and validation of a range of currently available detection methods developed from EWSs. We include an additional constraint, which requires multiple time-series points to satisfy the algorithms’ conditions before a detection of an approaching critical transition can be flagged. We apply this procedure to a simulated study of an infectious disease system undergoing disease elimination. For each detection algorithm we select the hyper-parameter which minimises classification errors using receiver operating characteristic (ROC) analysis. We consider the effect of time-series length on these results, finding that all algorithms become less accurate as the amount of data decreases. We compare EWS detection methods with alternate algorithms found from the change-point analysis literature and assess the suitability of using change-point analysis to detect abrupt changes in a system’s steady state.

## Introduction

Many attempts have been made to identify indicators of critical transitions in real systems. For infectious disease systems, criticality typically occurs when the basic reproduction number equals 1. Research has shown that infectious disease systems undergo the phenomenon of Critical Slowing Down (CSD) when the basic reproduction number, *R*_0_, crosses through the critical transition at *R*_0_ = 1 [1–3], reviewed in [4]. CSD describes the changes in the statistical properties of time-series data as a system approaches a critical transition, such as increased autocorrelation, variance and magnitude of fluctuations, due to the system’s slow recovery from perturbations as its dominant eigenvalue approaches zero. These statistical trends in time-series data serve as early warning signals (EWSs) of critical transitions. EWSs can be generated in real-time from time-series data and offer a computationally inexpensive technique for monitoring the progress towards elimination. Moreover EWSs are model-independent, in that they do not rely on empirically fitted models, and can be used to investigate approaching critical transitions in a wide range of real-world systems from ecological collapses [5–8] to climatic shifts [9–12].

The implementation of EWSs on empirical data will often use Kendall’s *τ* statistic to indicate the presence of a critical transition [1, 2, 13], researchers have reported that the observed trend of an EWS matches prior beliefs about when a bifurcation occurred in the system. However, research addressing *when* the trend of an EWS became significant, which we refer to as the “time-of-detection”, is limited. Reporting the time when an EWS gives a notable signal of an upcoming bifurcation will allow policy-makers to assess when a disease is close enough to elimination to die out without further intervention, thus prompting the end of a control campaign. The more advanced the warning, the more valuable EWSs will be in practice.

The field of change-point analysis is closely linked to the field of EWSs. Change-point analysis relates to identifying when the probability distribution of a time-series changes. Change-point analysis is used to identify when any abrupt change occurs and provides methods designed to detect the timing of such event(s). The first work in change-point analysis was in the 1950’s [14, 15], motivated by designing a method to automatically detect failures for quality control in manufacturing. This is a classical problem for detecting abrupt changes in the statistical behaviour of an observed signal, with extensive applications to climate modelling [16], network security [17] and fraud detection [18]. It is broken into two types of change-point analysis: offline, when the full data are available and we aim for the most accurate detection of the change-point; and online, for real-time monitoring of change-points. EWSs attempt to infer a single change point, where the system is undergoing a bifurcation, using signatures of the CSD phenomenon [10, 19]. Manifestations of CSD include changes in the variance and autocorrelation of the fluctuations of time-series data as the system approaches a critical transition. Moreover, change-point analysis can detect changes in the variance or correlation, by monitoring the probability distribution describing the time-series data and predicting the most likely timepoint where the change occurs. Thus change-point analysis can be used find when a change in an EWS can be detected rather than detecting the transition itself.

These two areas, although studying related questions, have little overlap in the literature. In some cases change-point analysis has been dismissed in the field of EWSs as it does not offer an advanced detection of a bifurcation [20]. However, the offline approach still offers the potential for predicting when a system underwent a bifurcation, which many in the field of EWSs desire. Offline change-point methods can be used to test the performance of EWS based techniques [21–23] and can provide an upper limit to the diagnostic performance expected of a real-time detection method.

The only study which proposes using EWSs within a change-point framework is Carpenter *et al*., [24] who implement an online change-point analysis (called Quickest Detection) by detecting the statistical signatures of CSD. The Quickest Detection method calculates the likelihood ratio for each data point and the ratio is updated as each data point becomes available, this is known as the Shiryaev–Roberts (SR) Procedure [25, 26]. To be implemented it requires the users to specify two probability distributions which describe the data pre- and post-critical transition, as well as specifying the detection threshold [27]. Establishing suitable choices for probability distributions and threshold criterion can influence the result of the time-of-detection, limiting the generality of the method [24]. Unkel *et al*., [28] reviewed the use of the Shiryaev–Roberts procedure for the early detection of disease outbreaks. In this work, they were interested in methods which could detect “outbreaks of concern”, rather than methods which detect when a disease crossed through the critical transition at *R*_0_ = 1. In particular, we are interested if similar statistical methods can be used for the early detection of disease elimination by considering the generalised behaviour of dynamical systems on the approach to a critical transition.

We compare a simple offline change-point analysis approach using the maximum likelihood estimation with the Quickest Detection method and with three other methods found in the EWS literature. We combine knowledge of CSD with change-point analysis, to find the first time an upcoming bifurcation can be detected, applied to an infectious disease system approaching elimination. In particular, the three methods we evaluate for detecting a critical transition using EWSs are:

1. **Exceeding two standard-deviations (known as 2-sigma)**. Drake & Griffen [29] proposed that an EWS, or composite of EWSs, is significant if it exceeds its long-run mean plus two long-run standard-deviations. The time-of-detection is given by the first time the EWS exceeds this threshold.
2. **The time changing *p*-value score of Kendall’s** *τ* **statistic**. Harris *et al*. [30], evaluate the significance (*p*-value) of Kendall’s *τ* score of an EWS by using a bootstrapping method to create a null distribution. This procedure is applied to increasing lengths of data segments from the start of the time-series up to the critical transition and the time-of-detection is inferred from the first time the *p*-value drops below 0.01.
3. **Logistic composite measure**. Brett & Rohani [31], consider a weighted sum of EWSs, where the weights are assigned using logistic regression. This composite measure is said to be significant if it exceeds a threshold which is identified by minimising classification errors. The time-of-detection is given by the first time when the logistic measure exceeds the threshold.

Each method is designed for real-time implementation (e.g. an “online” framework), where the EWSs are updated as new data are observed and a detection is triggered when the threshold criteria is exceeded. These methods have previously been applied to empirical studies, where the time of the critical transition was known and thus they could report the “lead-time” of each EWS. The “lead-time” gives an indication of how early on (prior to the transition) a change in the system could be anticipated, quantifying the power of EWSs of critical transitions.

Evaluating the available methods found in the literature is needed to understand whether there is a high false alarm rate, which would limit the potential for integrating EWSs with public health systems. For disease elimination, it is crucial that features of EWSs are not detected in data that are not undergoing a bifurcation, as this may lead to disease controls being incorrectly lifted. Therefore a highly specific method is essential. However, it is not necessary to have a large lead-time; instead of being used to prevent a critical transition, EWSs could be used to confirm the path to elimination. In contrast, for disease emergence, it is preferred to identify all possible cases which could lead to an epidemic or pandemic even if they are incorrect (i.e. high sensitivity) and ideally a method which gives a large lead-time so that there is enough time for control measures to be implemented and alter the course of the disease.

We evaluate each method on the same simulated epidemiological time-series data, for synthetic data that are: (1) at steady state and unchanging, (2) approaching disease elimination but levels off just before and (3) undergoing disease elimination. The first two data types represent two different null datasets, where we do not want to identify a bifurcation, with the second null dataset being more realistic. The purpose of this study is to validate each detection method and compare their performance. Some of the methods we consider were initially presented for climate or ecological data and thus have previously been implemented on large datasets that are rarely available in epidemiology. A key limitation with informing disease elimination with empirical datasets is to understand how EWSs behave in data poor settings and address their suitability under these circumstances. Therefore, we test each method for different lengths of time-series data, to provide quantitative evidence for which method performs best for varying amounts of data.

The 2-sigma method has been previously adapted by Clements *et al*., who found that by monitoring when two consecutive timepoints crossed the threshold (rather than 1), the frequency of false positive detections significantly decreased from 13% to 7% [32]. We extend the idea of a consecutive point strategy to all of the online detection methods we consider. We present a receiver operating characteristic (ROC) analysis to evaluate the “best” number of consecutive points, which we define as the number of consecutive points which minimises the classification error. We validate each method using a single point and measure the increased sensitivity and specificity when a constraint on the number of consecutive points is considered. Using the consecutive signal approach, we compare the first time disease elimination can be detected and the lead-time for each detection method and discuss the suitability of detecting disease elimination with these methods.

## Methodology

Below we describe the three EWS based methods in detail, followed by the two change-point analysis based methods (summarised graphically in Fig. 1–4). The colours used in Fig. 1–4 highlight similarities between the different methods. Red shaded boxes describe the input of the time-series data; green shaded boxes describe the calculation of a composition of EWSs (if applicable) and blue shaded boxes present the calculation of the time-of-detection for each methodology. In addition, we have provided open-source Python code located at https://github.com/ersouthall/Time-of-detection, which can be used to implement the detection algorithms found in this section and reproduce all results in this paper.

**Fig 1.**
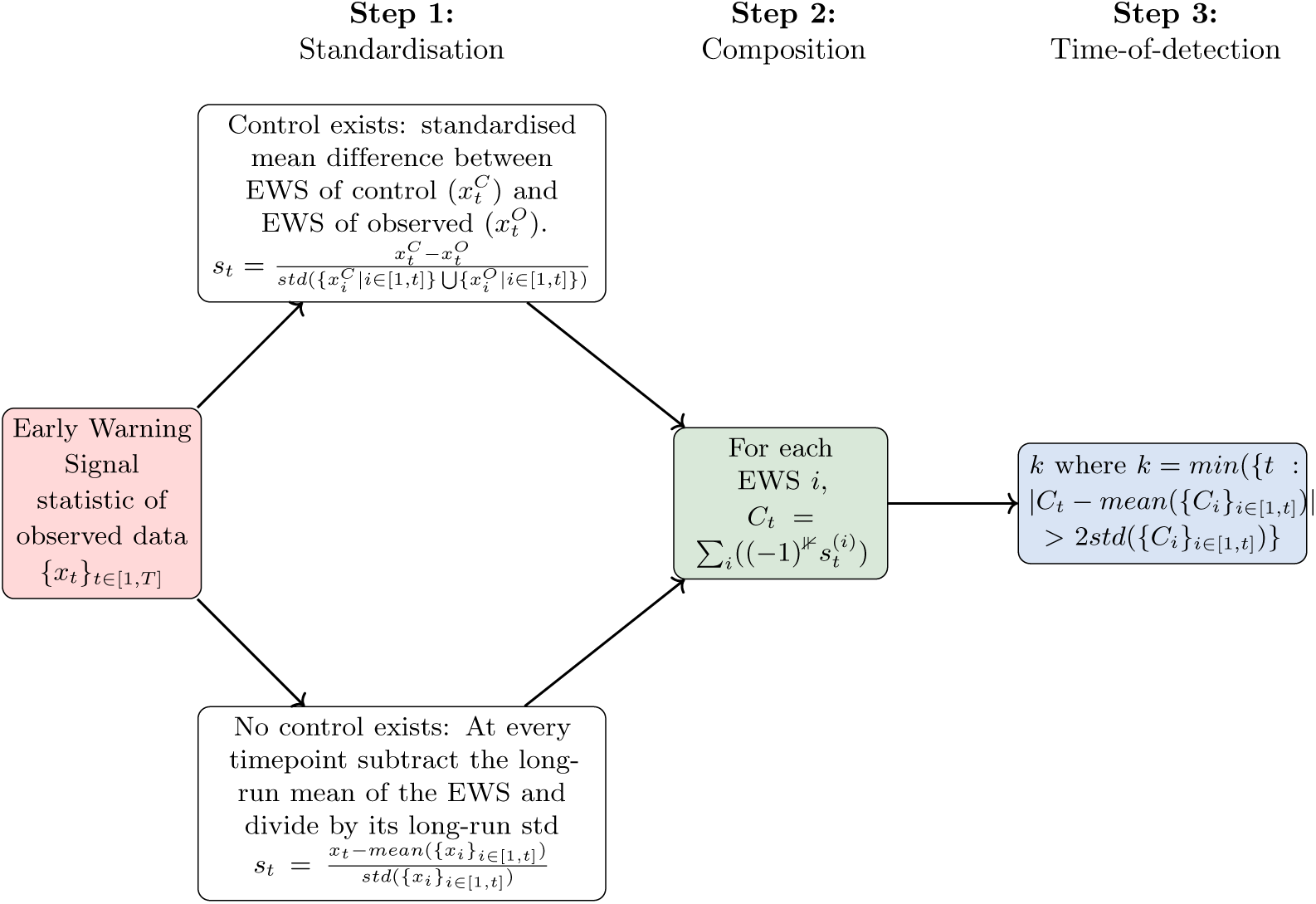
Flow diagram of the 2-sigma threshold method with the indicator extension from [33] (step 2, indicator function) – multiply the standardised EWS by −1 if the indicator is expected to decrease prior to the critical transition.

### Normalised Composite (2-sigma threshold)

Drake & Griffen first introduced the idea of anticipating a bifurcation with EWSs using a detection-based method [29]. They discussed taking a composition of normalised EWSs and detecting a critical transition when the composition crossed two standard-deviations (2*σ*). This method was initially applied in an experimental study, to detect the onset of environmental deterioration and offered a 110 day lead-time of the bifurcation [29]. Since, it has been implemented to a number of systems to detect: population collapse [33], recovery of a ecosystem [32] and the emergence of COVID-19 [34], although it has not been formally tested with a simulated control environment [35].

The 2-sigma method works by monitoring the long-run mean of a single EWS or a composition of multiple statistics. If a time-series is not going through a bifurcation, its value tends to revert to its long-run mean and properties of the time-series are not affected by the change in time [36]. Conversely, a non-stationary time-series (e.g. a time-series that is going through a bifurcation) does not tend to return to its long-run mean value and hence, its mean, variance and covariance will change over time. The 2-sigma method utilises this property of the long-run mean and defines the time-of-detection as the first timepoint when the EWS exceeds its long-run mean plus two times its long-run standard deviation.

In order to take the composition of multiple different EWSs, which might have different magnitudes, each EWS is standardised before calculating the sum *C*_*t*_ (Fig. 1). In particular, two different approaches are presented for standardising the time-series, depending on whether a control study exists (which is unlikely for epidemiological data).

An adaptation of the methodology was included in [33] using an indicator function which depends on the time-series trend of each EWS. In this research, taking the negation of EWSs which are expected to decline prior to the tipping point was found to result in a more robust method than taking the positive summation of all EWSs [33, 37]. The trend of an EWS can depend on the type of data used [3] and hence it is important to first analyse each complex system analytically to determine the trend of each EWS, before applying the indicator adaption.

### Changing *p*-value

A detection method based on significance testing of EWSs has been presented by Harris *et al*. [30]. In this work, the performance of EWSs are assessed in relation to their *p*-value score and the authors propose calculating a time-series of the changing *p*-value score for each EWS. This research highlights the issue of estimating the null distribution (in order to find the *p*-value) without access to replications. Here, a bootstrapping technique is proposed, which preserves the original features of the observed data and can be used to create the null distribution. However, the use of statistical tests and the misinterpretation of *p*-values have been previously criticised [4, 38]; and other approaches have been proposed for estimating the null distribution.

The method in Fig. 2 tries to infer if a system is going through a critical transition using a single EWS, by evaluating the *p*-value of Kendall’s *τ* score. As a system approaches a critical transition, Kendall’s *τ* score of some EWSs are close to the extremities (e.g. Kendall’s *τ* score is near 1 for CV and near −1 for variance, prior to disease elimination for EWSs calculated on incidence data [3]). This method is conditional on the null hypothesis, which states that there is no statistically significant relationship between Kendall’s *τ* score and whether the data are undergoing a bifurcation. The *p*-value is the probability of observing a more extreme Kendall’s *τ* score, conditional on the null hypothesis being true. For each bootstrapped time-series, Kendall’s *τ* score is calculated to create a null distribution of scores, which can be used to evaluate the *p*-value of Kendall’s *τ* score of the observed time-series.

**Fig 2.**
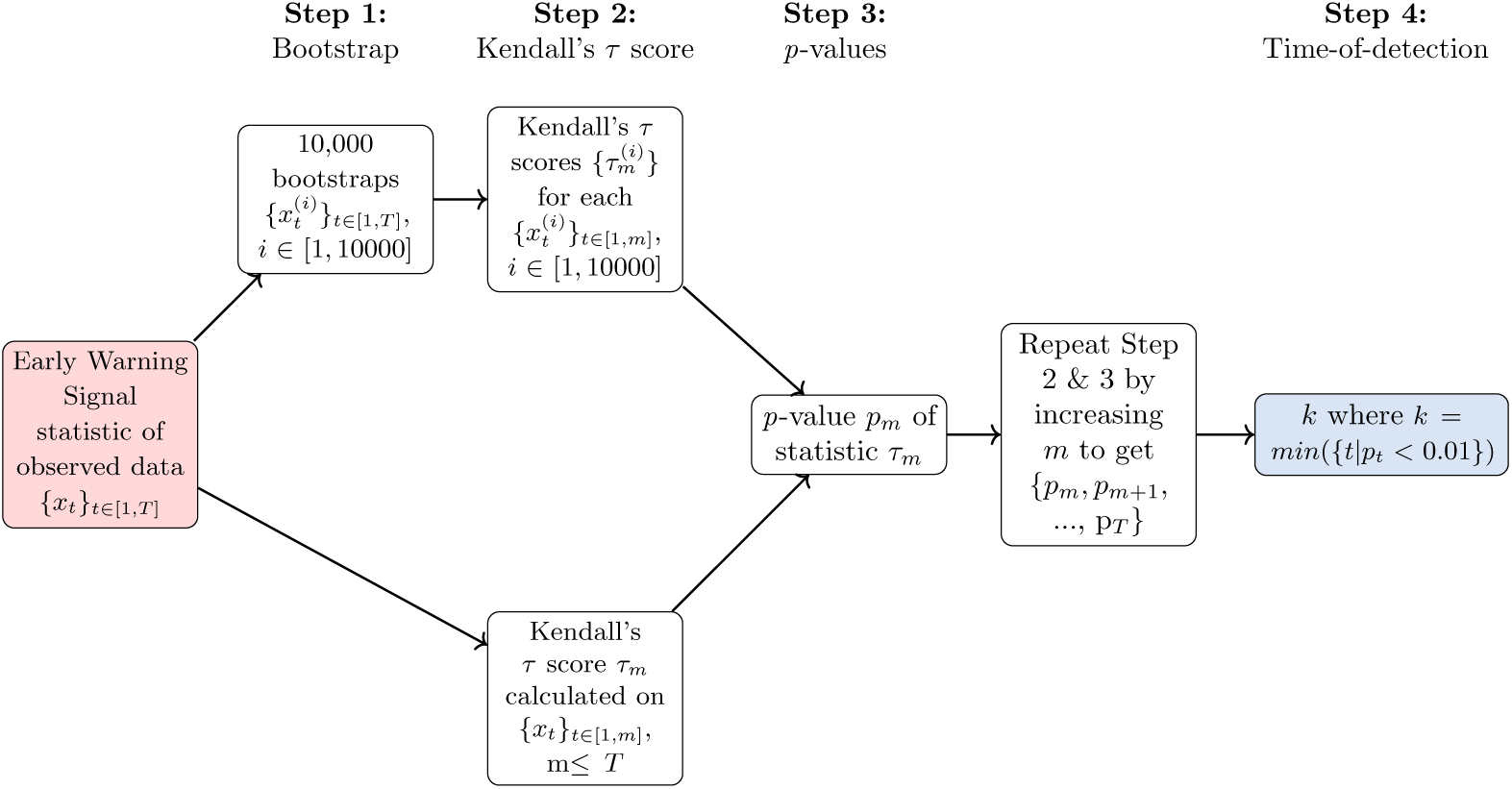
Flow diagram of the changing *p*-value detection method.

The methodology is implemented on a subsample {*x*_*t*_}_*t*∈[1,*m*]_, *m < T* of the observed time-series. The length of time-series used to calculate the *p*-value is increased (i.e. by increasing *m*), to create a time-series of *p*-values, where each *p*-value is calculated with more complete information of the entire time-series {*x*_*t*_}_*t*∈[1,*T*]_. The point at which the *p*-value crosses below 0.01, is recorded as the time-of-detection.

### Logistic Transform Risk

Clements *et al*., [39] reviewed the use of multivariate statistics in EWS literature and noted that a more sophisticated analysis, such as the use of machine learning algorithms with composite statistics would be an obvious expansion. This was subsequently explored by Brett & Rohani [31]. They computed a logistic regression of a weighted sum of EWSs, where the weights were determined using a lasso regression with a cross validation technique.

This method can be implemented on a single EWS or on a composition of multiple EWSs. A supervised machine learning algorithm is implemented (outlined in the pre-steps in Fig. 3) on simulated data to train the algorithm’s parameters. In particular, simulations are run under two scenarios: a fixed system (controlled environment) and a system undergoing a bifurcation (changing environment). These simulations are then labelled as controlled and changing; and the logistic transform method can be tested on its ability to correctly classify the simulations. In particular, for each EWS *i*, a binary logistic regression with lasso regression (*l*^1^−penalty) is used to get the optimum weights 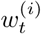 and the intercept *w*_0_, where the training data are used for refining the weights and training the algorithm. This assumes that the timepoints for each EWS time-series is an independent data point and assigns equal importance to all data points when fitting the logistic regression.

**Fig 3.**
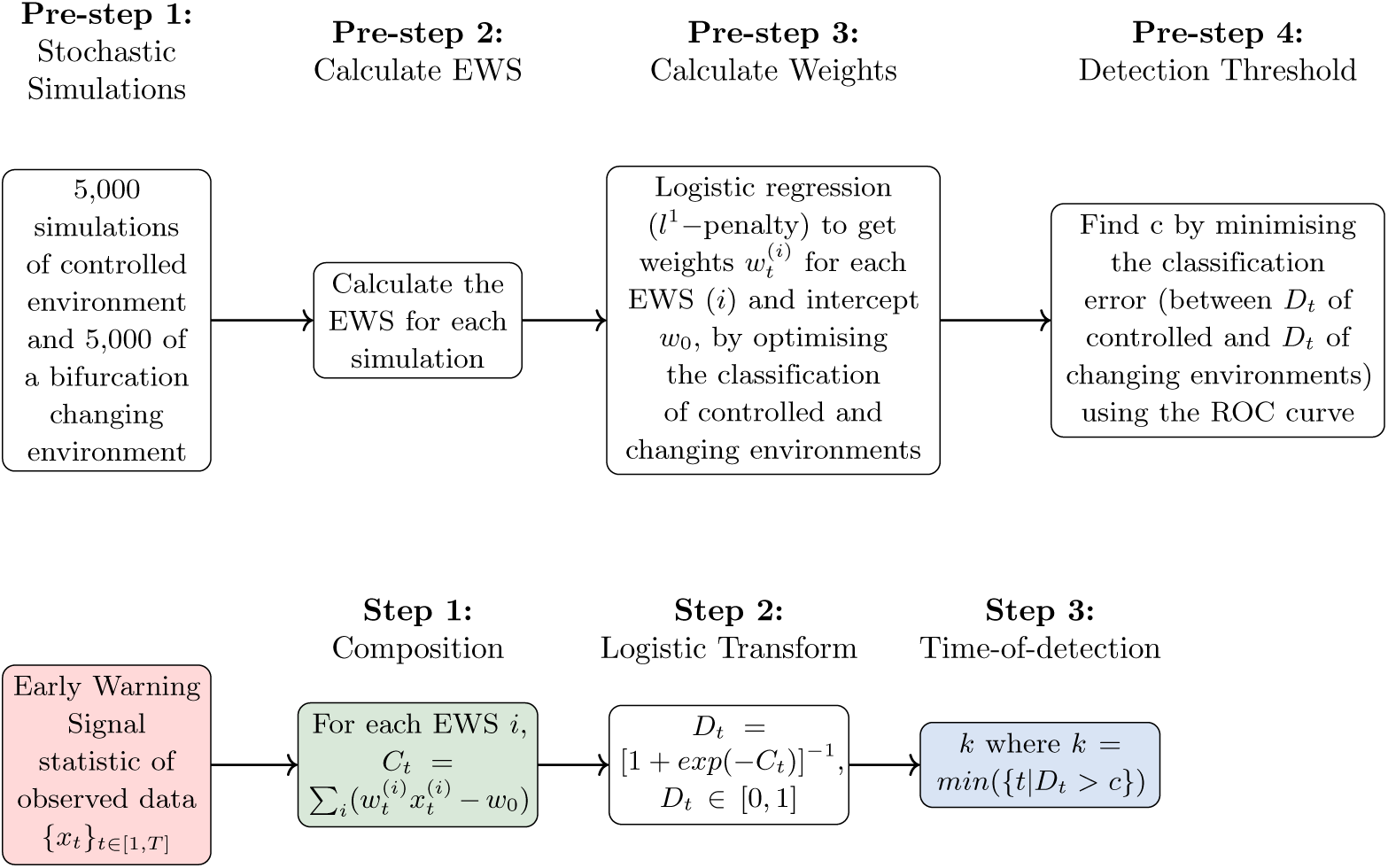
Flow diagram of the logistic transform detection threshold method.

**Fig 4.**
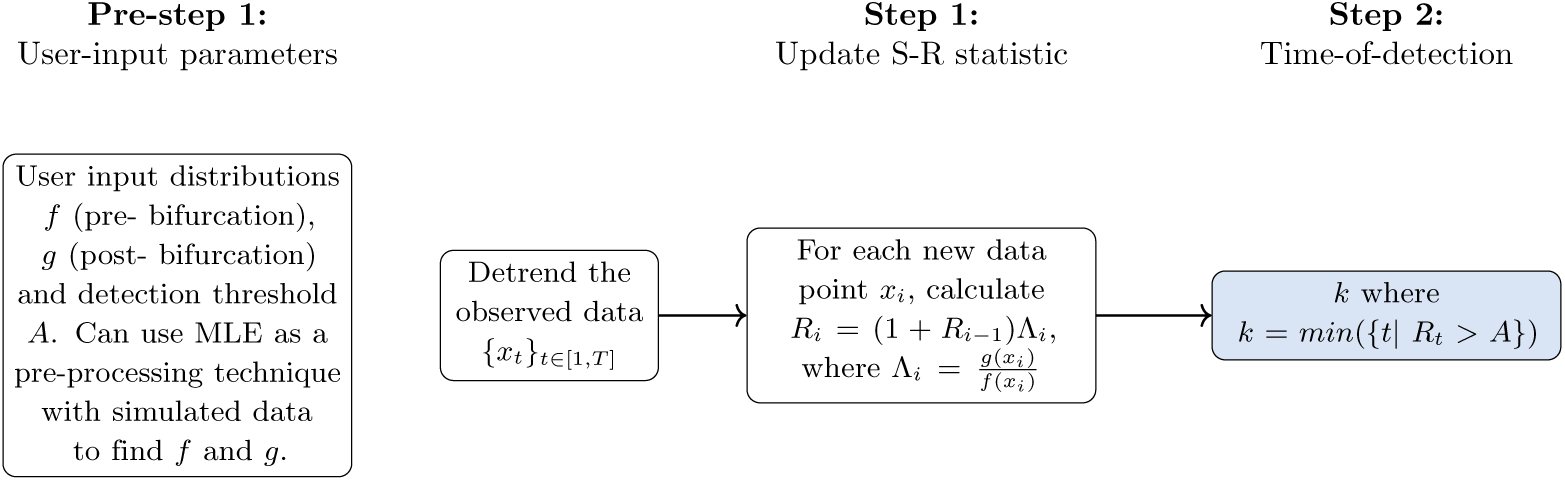
Flow diagram of the Quickest Detection method.

A logistic transformation of a weighted composition of EWSs, *D*_*t*_, is considered and the time-of-detection is inferred when *D*_*t*_ *> c*, where *c* is the detection threshold. The value of *c* is also found using a supervised learning technique, by computing the ROC curves and evaluating the value of *c* which minimises the false positive rate and maximises the true positive rate. Once the value of *c* is chosen during training, it is then used with the observed data (step 1 to 3 in Fig. 3).

We adapt the logistic regression method to use normalised training data rather than raw data. We find that when testing the logistic transform method with data from different settings, such as with a lower population size or a smaller initial number of infections, the weights are not appropriately scaled for the testing data. Instead, we normalise the training data using a z-score standardisation of EWSs, equivalent to Drake & Griffen’s long-run standardisation (see Fig. 1). A standardisation of EWSs allows the weights to be independent of the data scale and greatly improves the generality of the logistic transform method.

We generate the training dataset using the same SIS dynamics as the test dataset (described in Supplementary Methods S1). While, Brett & Rohani implemented their method for disease emergence [31], they demonstrated that the same weights could be used for the emergence of pertussis, mumps and even plague. Here, by evaluating the obtained weights with a training dataset which has the same SIS dynamics as the testing dataset, we are assessing the best predictive performance of the logistic classifier. We only train the classifier using data of length 100, which assumes that the weights will be independent of sampling or length of the time-series. Additionally, we calculate the EWSs using a moving window of 50 timepoints for the training dataset and 30 for the testing dataset, to reduce overfitting.

Our training data consists of 10,000 Gillespie simulations of incidence data; comprising of 5,000 simulations of a disease approaching elimination (bifurcation changing environment, named Ext) and 5,000 simulations of a disease at the endemic steady state (controlled environment, named Fix). We implement the logistic method using the EWSs: autocorrelation lag-1; autocovariance; standard deviation; skewness; kurtosis; coefficient of variation and index of dispersion. We reduce the total number of combinations of these seven EWSs from 127 to 17, by removing combinations of EWSs where the predicted coefficients are zero and by only considering combinations of EWSs which achieved an *AUC ≥* 0.6 with the training data. The weights of each EWS depend on the combination of EWSs, but in general the autocovariance and the standard deviation are negative; while AC(1), skewness, kurtosis and coefficient of variation are positive. The index of dispersion was the only EWS to sometimes be negative and at other times be positive.

## Change-point Analysis Methods

### Maximum Likelihood Estimation

Maximum likelihood estimation (MLE) was first proposed by R. A. Fisher in 1922 [40]; it is a popular method for approximating unknown parameter values that describe or fit to some observed data. It works by assigning a likelihood ℒ (Θ|*X*) distribution, describing the probability of the parameters of the system being Θ giving the data, *X*.

The MLE approach can also be used to find the optimal timepoint for when the probability distribution describing the data changes. Analytically, it has been shown that the fluctuations about the steady state can be described with a Gaussian distribution with a mean of zero and a variance [1, 3]; thus we use the Gaussian distribution as the likelihood for the MLE method. We aim to find the optimal timepoint in the data describing when the variance of the fluctuations changes. Under this framework, the data prior to change point, *τ*, is described by a Gaussian distribution with variance 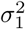 such that 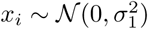 for *i* = 1, …, *τ* and after the change point the underlying probability distribution has a variance 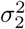 e.g. 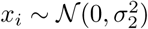 for *i* = *τ* + 1, …, *T*. Our null hypothesis assumes that the two variances (and distributions) are equal, 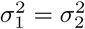 and the alternative hypothesis is that a single change point *τ* exists such that *σ*_1_ ≠ *σ*_2_.

Therefore the null hypothesis (*H*_0_) is given by,

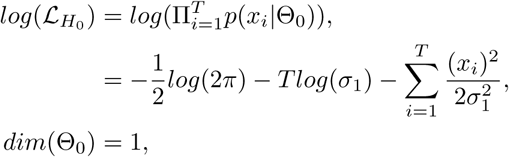

and the alternate hypothesis (*H*_1_),

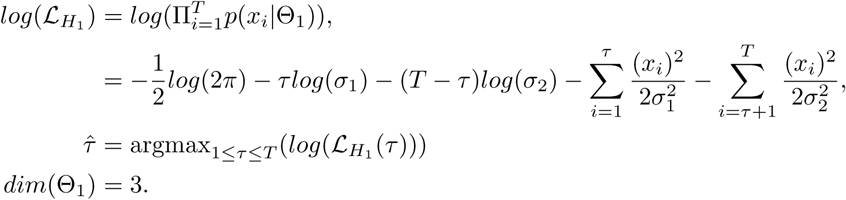

We require the variance to satisfy *σ*_1_ ≥ *σ*_2_ ≥ 0 for incidence-type data (e.g. the variance of the fluctuations is decreasing and positive, as demonstrated analytically in [3]) and *σ*_2_ ≥ *σ*_1_ ≥ 0 for prevalence-type data (e.g. the variance is increasing, see [3]). Compared to the other detection algorithms we analyse, the MLE technique is performed on the entire time-series, rather than in real-time. Additionally, this method does not consider that the variance may be changing gradually and so it is perhaps more useful for examining if a bifurcation has already occurred in the system, rather than anticipating a future bifurcation. The change-point detection is based on the likelihood ratio procedure using model selection penalties. Hypothesis testing is performed with the likelihood ratio test, 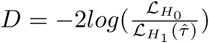 and a penalty criteria, to decide whether the change-point *τ* is significant or not. If the null hypothesis is rejected then the value of 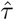 is accepted. In the hypothesis testing, we compare the likelihood of Θ ∈ Θ_0_ with Θ ∈ Θ_1_ and often the model selection criteria depends on the property *k* = *dim*(Θ_1_) − *dim*(Θ_0_) = 2. Different penalty criteria can be considered to decide where to reject the hypothesis and usually the null hypothesis is rejected when *D > λ*, where *λ* can be defined as:

- **No model selection:** Accept all, *λ* = 0,
- **Hannan-Quinn information criterion** (see [41]): *λ* = 2*klog*(*log*(*T*)),
- **Akaike information criterion (AIC)** (see [42]): *λ* = 2*k* = 4,
- **Wilks’ Theorem** (see [43]): At the 5% level, 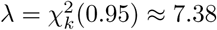,
- **Bayesian information criterion (BIC)** (see [44]): *λ* = *klog*(*T*),
- **Modified BIC (MBIC)** (see [45]): *λ* = *log*(*T*) + *log*(*τ*) + *log*(*T* − *τ* + 1).

Without model selection, the null hypothesis will always be rejected, thus when implementing the MLE approach on a stationary time-series the percentage of false alarms will be 100%. As *λ* is increased, the specificity of the algorithm will increase, although the sensitivity could decrease. Hannan-Quinn and the two BIC criteria depend on the length of the time-series *T*, and for the lengths we examine, the criteria for Hannan-Quinn is always lower than AIC (*λ <* 4). The BIC criteria is sometimes lower than Wilks’ Theorem (e.g. at *T* = 20) and other times greater and the MBIC criteria will always be the greatest unless the value of *τ* = 0. An important property of the MLE states that 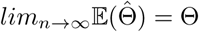, which implies that as the number of samples are increased, the estimated parameter values are closer to the truth. Thus, we expect the MLE to be more robust for longer lengths of time-series data.

### Quickest Detection

Quickest detection is an online approach which assumes that the time-series {*x*_1_, *x*_2_, …, *x*_*T*_} is drawn from a probability distribution *f* (*x*_*i*_) for observation *x*_*i*_ and that after a signal is detected, the time-series is described by the probability distribution *g*(*x*_*i*_) (see schematic 4). A detection is triggered when the likelihood ratio of *g*(*x*_*i*_) and *f* (*x*_*i*_) exceeds the threshold *A*. The likelihood ratio, also known as the Shiryaev–Roberts (SR) Procedure (see [25]), is given by the recursive relation *R*_*n*_ = (1 + *R*_*n*−1_)Λ_*n*_, *R*_0_ = 0, where 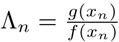. Like the EWS based methods for time-of-detection, the quickest detection method is updated every time a new data point arrives, making it suitable to be an automated process. The change point *τ*, is the first time when *R*_*τ*_ *> A*.

The quickest detection method requires the user to choose useful distributions for *f, g* as well as the value of *A*. The threshold, *A*, can be described at the expected number of time steps to a false positive given a tolerance to false alarms. The method can be sensitive to the choice of *f, g* and *A*. Carpenter *et al*. [24], suggest fitting a Normal distribution *f* to the first 50 timepoints of the time-series and then describing *g* with a Normal distribution with a larger mean and variance, chosen from tests on simulated time-series. By implementing the MLE method first, distributions for *f* (*x*) and *g*(*x*) can be found using the entire time-series of a single simulated training dataset and then the quickest detection method can be tested in real-time on the testing datasets.

### Evaluating Detection methods

To compare each method, we assess its ability to detect a signal of an approaching disease elimination critical transition and examine how early the critical transition can be detected. We also assess each method’s ability to not detect any critical transition in a disease process where a transition is not present. We use the same Gillespie simulation study of incidence data from Southall *et al*. [3] (further details are provided in Supplementary Methods S1), involving 500 simulations of the SIS model going through a bifurcation (i.e. going extinct, named Ext), 500 simulations of the SIS model at steady state (named Fix) and 500 simulations of the SIS model which have the same initial dynamics at Ext however do not reach elimination (i.e. not going extinct, named NExt). The data and code used to run these simulations can be found at https://github.com/ersouthall/Time-of-detection.

We evaluate each detection method using the same testing data of Ext, Fix and NExt simulations. We sample the original time-series (of length 500) and consider how the detection methods perform for different lengths of available data. In particular, we consider four different lengths of time-series: 20 timepoints, 50 timepoints, 100 timepoints and 250 timepoints. The study is designed so that the bifurcation should occur at 80% *× length*(time-series) and we measure the lead-time between a detected signal and this timepoint.

#### Sensitivity, specificity and the power metric

We compare the sensitivity and specificity of each method and between EWSs. The power metric [46] combines the score of sensitivity and specificity into a single performance measure and can be used to evaluate the predictive power of each method for each EWS. The power metric is defined as,

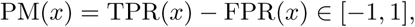

where *x* represents a detection method and TPR is the true positive rate (e.g. out of the 500 simulations of Ext, the proportion which detect a critical transition) and FPR is the false positive rate. A score closest to 1 is the best and a score close to −1 is the worst. Using the power metric allows us to rank indicators and composition of indicators by maximising both sensitivity and specificity.

As we are considering two null datasets (Fix and NExt data), we define the false positive rate of Fix simulations as FPR_1_ and of NExt simulations as FPR_2_. Similarly, the true negative rate (TNR) is given as TNR_1_ and TNR_2_ respectively. To account for two null datasets, we adjust the power metric giving,

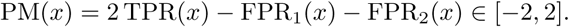

#### Extension: consecutive timepoints

In each method discussed above, the time-of-detection is given by the first timepoint when the metric considered exceeds the defined threshold. We modify this definition, by taking the time-of-detection to be the first time when multiple consecutive points are above the threshold. This constraint removes scenarios when an anomalous timepoint, or isolated timepoints, are observed above the threshold but the majority lie below. As a system approaches a critical transition, we expect the number of consecutive points above the threshold to increase, which would more likely reflect a critical transition. Using multiple consecutive timepoints will also help to highlight denser regions which lie above the threshold, thus giving a stronger confidence of the lead-time.

A consecutive signal approach has previously been suggested with the 2-sigma composite method and evidence showed that using only two consecutive points is sufficient for reducing the number of false identifications [32, 37]. However, an analysis on the “best” number of consecutive points has not been conducted. In particular, the most suitable number of consecutive points may depend on the length of the time-series; the EWSs considered; the time-series data available (e.g. incidence or prevalence); the system’s dynamics (e.g. rate of elimination) and the detection method used.

We extend the proposal of using multiple consecutive points to examine the optimal number of consecutive points, which we find by minimising classification errors for each detection method. We examine the consecutive signal approach for all methods which use a threshold: 2-sigma composite, changing *p*-value, logistic transform and quickest detection. Error rates can be visualised by ROC curves [47]. ROC curves are constructed by measuring the sensitivity and specificity of each EWS at different threshold values. To minimise classification errors, we present discrete ROC curves for each EWS and find the threshold (number of consecutive points) which maximises the power metric from these curves. The performance of each detection algorithm is measured by the area under the curve (AUC) of the ROC curve. An AUC score of 1 will indicate that the algorithm correctly detected an EWS in all Ext simulations, and that there was also perfect specificity (no false detections were reported in the null datasets). We study the ratio of consecutive points of the dataset and whether there is a general trend in the best number of consecutive points by percentage of the time-series length. In particular, we consider if any significant issues arise when implementing the methods on shorter time-series.

## Results

In Fig. 5, we demonstrate how the online detection methods (2-sigma, changing *p*-value, logistic transform and quickest detection) work for a single time-series. Each detection method was initially proposed in the literature as detecting a transition when a single time-point exceeded the threshold (given by a single bold marker). In the example shown in Fig. 5, all methods result in some false alarms, whereby the null datasets, Fix and NExt, are shown to trigger a detection (green and purple bold markers respectively). However, for the Fix data these are sparse (e.g for the 2-sigma method Fig. 5(a)) and for the NExt data these are often in small clusters (e.g. for the 2-sigma and logistic composite method Fig. 5(a,c)). This supports the use of requiring multiple consecutive points to exceed the threshold, in order to trigger a detection. For a time-series which is undergoing disease elimination (Ext data, blue line), the concentration of detected points increases on the approach to the transition, until all points exceed the threshold. By implementing a consecutive point strategy, it will result in a later time-of-detection (smaller lead-time), since the earlier detected points (Ext data, blue) which exceeded the threshold will no longer satisfy the additional consecutive point threshold. Thus, when determining how many consecutive points are sufficient, there is a trade off between whether a high specificity is necessary or a longer lead-time.

**Fig 5.**
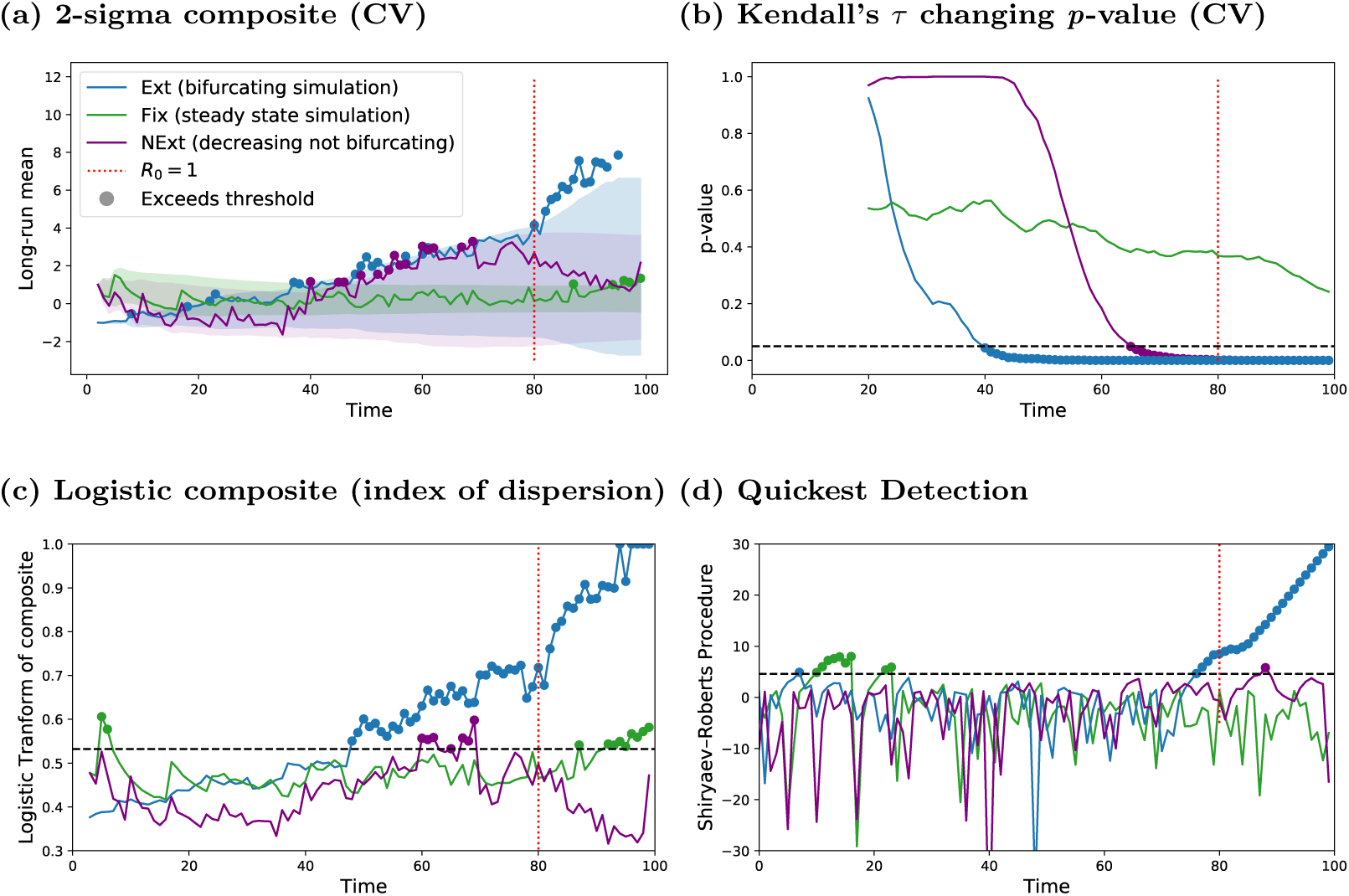
Comparison of detection methods: For an incidence time-series of length 100, each detection method is demonstrated on a single time-series of Ext data (blue line), Fix data (green line) and NExt data (purple line). The detection method considered: (a) 2-sigma composite framework: shown for a single EWS (CV), (b) Kendall’s *τ p*-value methodology: shown for a single EWS (CV), (c) the logistic composite methodology: shown for a single weighted EWS (InD) and (d) the quickest detection approach: shown with *σ*_1_ = 34, *σ*_2_ = 2.4 and *A* = *log*(*T*). Each detection method was initially described by detecting a critical transition when the statistic crosses the relative threshold; this is shown in these figures using bold markers. Blue markers highlight true classification and green or purple markers indicate false classification.

The exception is the changing *p*-value method, where nearly all 500 simulation sets exhibited a very high false detection rate with the NExt data (Fig. 5(b)). This method uses Kendall’s *τ* score and previous work by Dessavre *et al*. has reported the poor predictive power when classifying Ext simulations from NExt simulations using Kendall’s *τ* score [48]. In some cases, we find that when implementing the changing *p*-value method, the NExt data exceeded the threshold earlier and with more dense regions of detected points than the Ext data.

Clement *et al*., [33] recommended the use of two consecutive points with the 2-sigma method. However, we observe that in this example (Fig. 5(a)) a two consecutive point constraint is not sufficient to remove the isolated cluster of falsely detected NExt data points (purple bold markers). However, a two consecutive point constraint will remove all the anomalous detection points from the Fix data (green bold markers). Hence, by implementing the 2-sigma method on a more realistic null dataset of declining incidence, we find that a stricter number of consecutive points are needed. We propose to implement a similar consecutive point approach with the other online detection methods (Fig. 5(b-d)) and use ROC analysis to find the optimal number of consecutive points.

The power metric measures the difference between true positives and false positives (so that higher values represent a better ratio of true positives to false positives). We expect the power metric to initially increase as the number of consecutive points increases, as this will reflect a reduction in the number of false positive detected points without influencing the true positive rate. In Fig. 6, we present the results from our power metric analysis applied to multiple EWSs (full list of EWSs can be found in Supplementary Methods S2) for a time-series of length 100 and we investigate how the power metric changes when the number of consecutive points are increased from one (Fig. 6). In the Supporting Information we include the power metric analysis for time-series of length 20, 50 and 250 in Fig. S1. In Fig. 6 and Fig. S1, we show that in general the performance of all online detection methods improve with the consecutive points. The quickest detection method is the best method for maximising the power metric for all time-series lengths and is the only method to return a reasonable power metric score when a single consecutive point is used (left most column in Fig. 6(a-d)). In general, a suitable detection threshold *A* for the quickest detection method can be described as *A* = *log*(*T*), where *T* is the length of the time-series.

**Fig 6.**
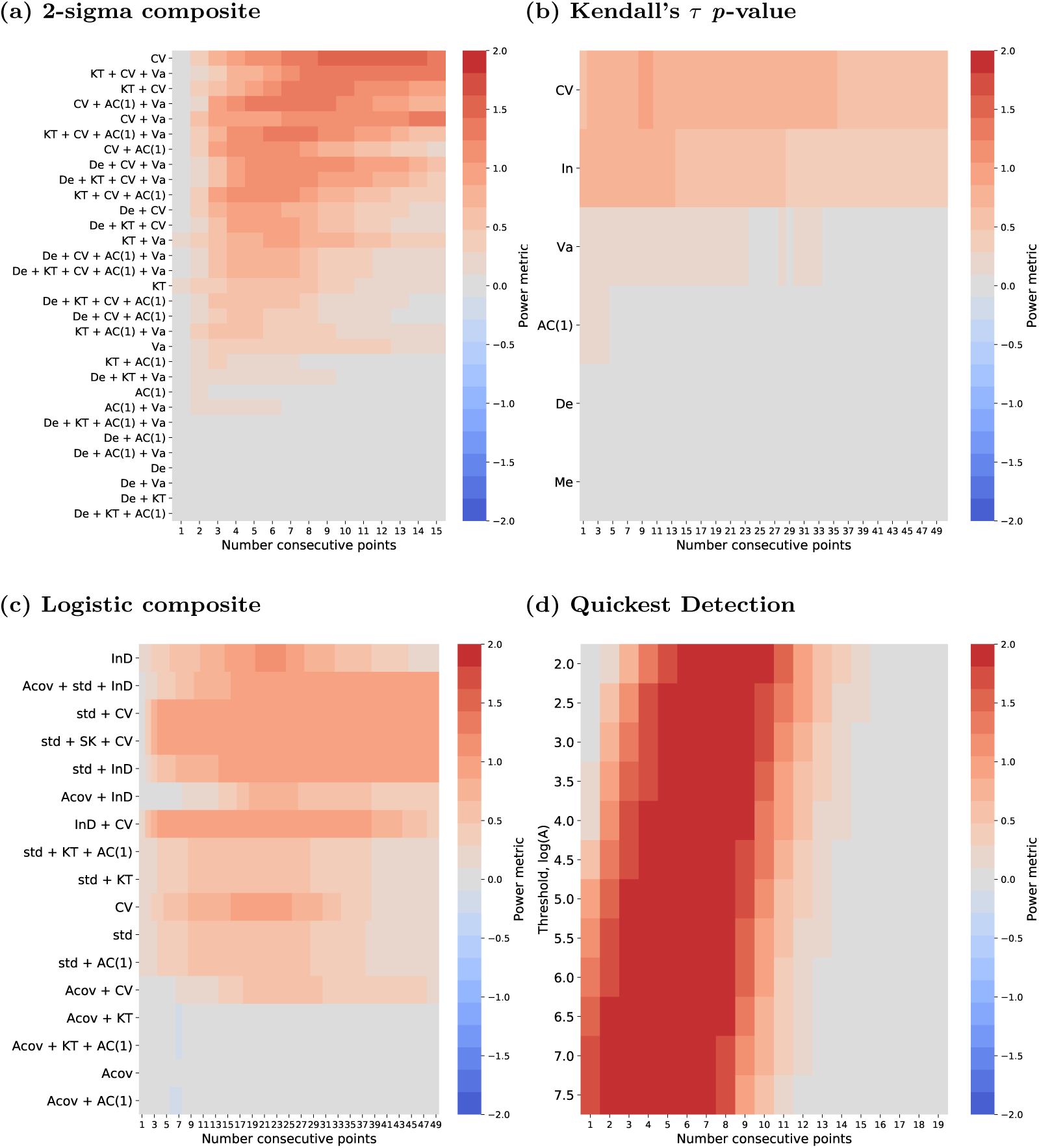
Power Metric Analysis: For a time-series of length *T* = 100, the power metric is calculated as 2*TPR* − *FPR*_1_ − *FPR*_2_∈ [−2, 2] for each statistic. The power metric results are shown for each online detection method considered: (a) 2-sigma composite, (b) Kendall’s *τ p*-value, (c) the logistic composite and (d) the quickest detection approach. Each detection method was initially described when the number of consecutive points was one (left column in all panels). Each figure demonstrates how the power metric changes when a stricter constraint on the number of consecutive points are required to cross the threshold before the detection is triggered. A list of EWSs and their acronyms used in this figure can be found in Supplementary Methods S2

Although the quickest detection method performs best when comparing the power metric, we show that when only the first 80% of the time-series is analysed (i.e. data prior to the critical transition at *t*^*^ = 80) the power metric drops and the quickest detection method is no longer suitable (Fig. 7(b)). The power metric reduces for all detection methods when only considering data up to the bifurcation point (see Fig. S2), however this is the most severe for the quickest detection. This implies that the time-of-detection for the quickest detection method is delayed (post bifurcation) for nearly all 500 simulation sets and that the method is detecting disease elimination rather than an advanced indication that *R*_0_ is crossing through 1 due to a change in the variance. As such, there is little lead-time with this method, as it does not provide an early signal of the bifurcation. In comparison, the 2-sigma method also worsens (although more mildly) suggesting that the time-of-detection for some simulations occur after the bifurcation point. The logistic composite is the least impacted, implying that the logistic composite method gives the largest lead-time. In particular, with the 2-sigma method, we find that the composite CV+Va has a fairly strong power metric, although it is not as robust at CV (Fig. 6(a)), however when using just 80% of the time-series, CV+Va outperforms the other composites (Fig. 7(a)). This is due to the composite, CV+Va, obtaining the lowest FPR_1_ score. As, NExt and Ext data are the same up to *t* = 74, all composites struggle to minimise FPR_2_ prior to *t* = 80 and thus the power metric presented in Fig. 7 is driven by the FPR_1_ score.

**Fig 7.**
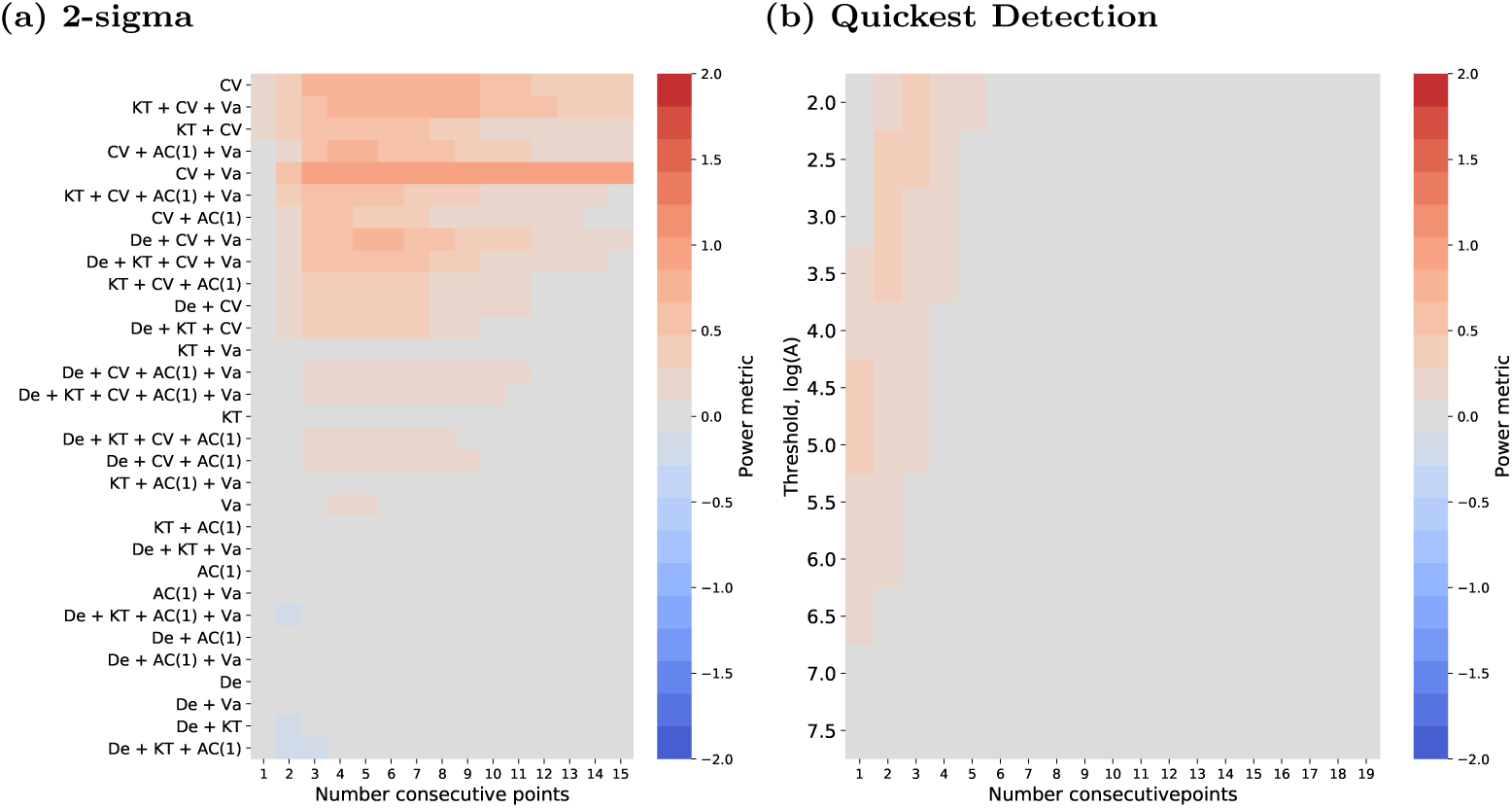
Power metric for time-series data up to the bifurcation point. For a time-series of length 100 and bifurcation occurring at *t*^*^ = 80, the total power metric (2*TPR* − *FPR*_1_ − *FPR*_2_), is calculated over the time-series data up to the bifurcation. Results are shown for each detection method considered: (a, 2-sigma composite) and (d, quickest detection). A list of EWSs and their acronyms used in this figure can be found in Supplementary Methods S2

The power metric can also be used to identify the worst statistics (score *≤* 0) and this information can be summarised by measuring the AUC score, as shown in Fig. 8. In Fig. 8, we illustrate the AUC describing the classification errors between Ext and Fix data (lower triangles) and for classification errors between Ext and NExt data (upper triangles). The AUC is calculated from ROC curves which illustrate how the sensitivity and specificity changes as the number of consecutive points above the threshold are varied. The optimal number of consecutive points can be found by selecting the best possible cut-off value from the ROC curves (i.e. the cut-off value which assigns the ROC curve point in the upper left corner). This is shown for a range of time-series lengths (time-series of lengths: 250, 100, 50 and 20) which we generate by sampling the incidence time-series data at different frequencies. We find that combinations of AC(1), decay time and variance perform poorly for all time-series lengths with the 2-sigma method (Fig. 8(a)). This is reflected with an AUC score near 0.5, describing a random classification between Ext simulations and the null datasets. A similar behaviour is observed when considering AC(1) and decay time with the changing *p*-value method (Fig. 8(b)) and combinations of AC(1) and autocovariance with the logistic composite method (Fig. 8(c)).

**Fig 8.**
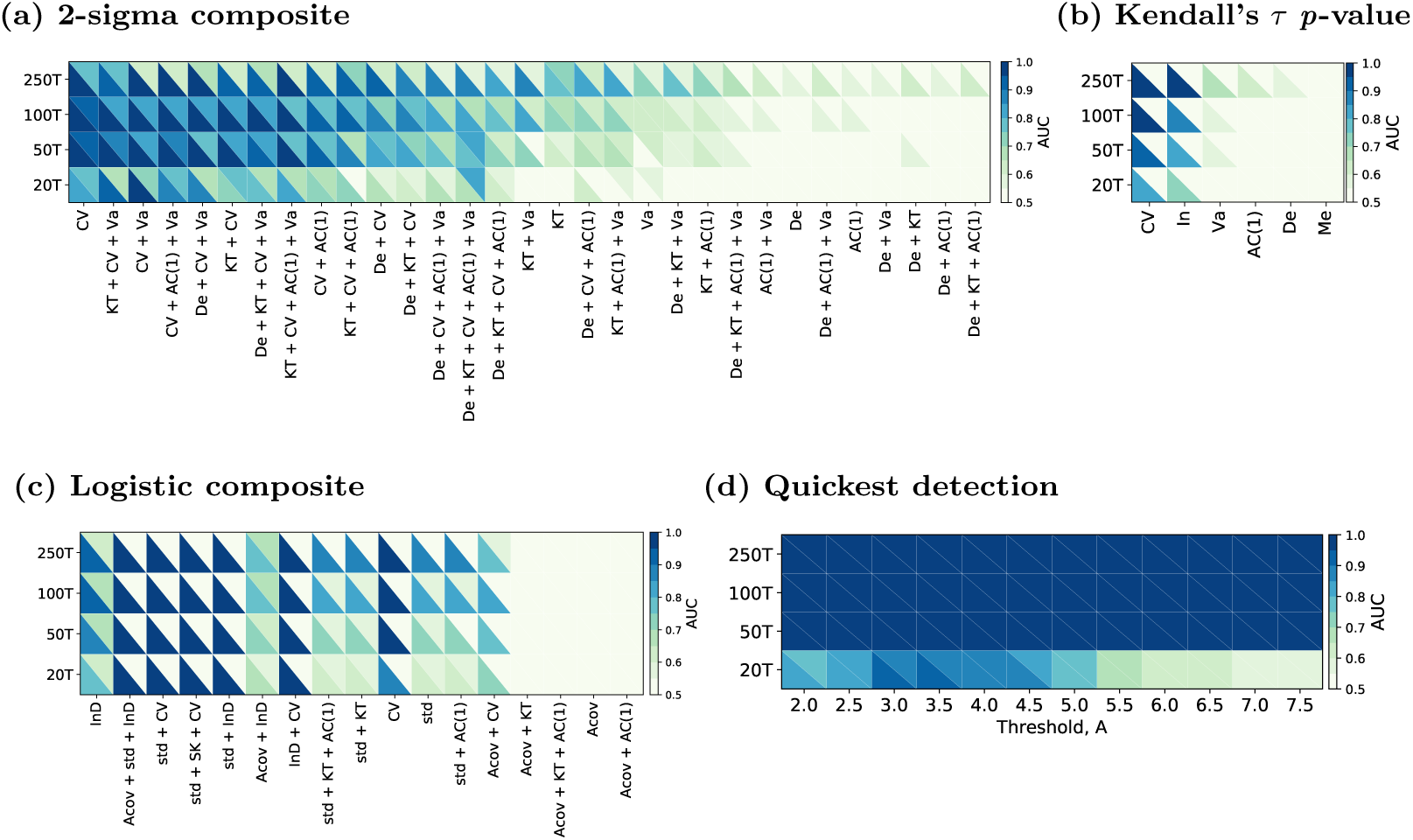
ROC Curve analysis (AUC): For each detection method, ROC curves are calculated for each EWS by recording the TPR and FPR scores for each constraint on the number of consecutive points required to exceed the detection threshold. The AUC of the ROC curve results are shown for: (a, 2-sigma method); (b, Kendall’s *τ* method); (c, logistic composite method) and (d, quickest detection method), indicating the predictive performance of each EWS for different length time-series considered (lengths: 250, 100, 50 and 20). Each heatmap shows the ROC curve analysis for the TPR given by disease elimination simulations and the FPR given by two different null models. The bottom triangles show the steady state null model (Fix) and the top triangles show decreasing incidence null model (NExt). A list of EWSs and their acronyms used in this figure can be found in Supplementary Methods S2

An ideal detection method and EWS, would have a robust AUC score (AUC ≥ 0.75) using both null datasets. In Fig. 8 this would correspond to blue shading in both the lower and upper triangles, which can be found for the quickest detection method (Fig. 8(d)) for all values of *A*, except for the shortest time-series, where it only holds for 2.5 *≤ A ≤* 4.5. Whereas, Kendall’s *τ* changing *p*-value is not a suitable method, as the AUC with NExt simulations is near 0.5 for all EWSs and all time-series lengths. For the 2-sigma method, the best combination of EWSs is CV which has an AUC score of (*AUC*_*F ix*_, *AUC*_*NExt*_) ≥ (0.99, 0.77) for all time-series lengths. In general, for the logistic composite method, combinations which contain index of dispersion (InD) perform well and similarly to the 2-sigma method std + CV is a suitable combination. The combination InD + Acov provides the highest AUC score with NExt simulation (*AUC*_*F ix*_, *AUC*_*NExt*_) = (0.75, 0.68), although InD gives the highest combined AUC score (*AUC*_*F ix*_, *AUC*_*NExt*_) = (0.92, 0.66) — neither combination are robust for time-series length 20.

We find the best number of consecutive points for each online detection method and for each EWS and we evaluate this as a percentage of the length of the time-series. This is summarised across all EWSs for each detection method in Fig. S3. In general, we observe that for quickest detection, the best number of consecutive points is less than 5% of the time-series and that for both null datasets, quickest detection achieves an *AUC ≥* 0.9 except for a time-series of length 20 (Fig. S3(a), 0.55 *≤ AUC ≤* 0.9) – agreeing with Fig. 8 and Fig. 6. Similarly, for the 2-sigma composite method, there is a clear general trend with the best number of consecutive points being between 5 − 10% of the time-series; however as found in Fig. 8, the AUC score is less concentrated and some compositions perform poorly. For the logistic method a higher number of consecutive points are optimal, with the general trend suggesting between 10 − 25% of the time-series length. However, some composites perform best with over 30%. There is not a general trend across time-series lengths for Kendall’s *τ p*-value method, with results ranging from 1 − 50%. As such, the biggest disadvantage of this method is that a large number of points are required to cross the threshold in order to minimise the classification errors. This is an impractical constraint and limits the potential of this method to be used in real-time.

We consider the EWS (or composite of EWSs) which achieves the highest AUC for each method and compute the time-of-detection of each simulation set. We evaluate the time-of-detection using the consecutive points strategy, where we use the best number of consecutive points for each EWS which we found during the ROC analysis. In particular, we examine the CV using 12 consecutive points with the 2-sigma method; the CV using 5 consecutive points with Kendall’s *τ p*-value method; the InD using 21 consecutive points with the logistic composite method and the quickest detection method with *A* = *log*(100) *≈* 4.5 using 6 consecutive points. Additionally, we implemented the MLE method, which is a retrospective method and cannot be considered in real-time.

We find that with the 2-sigma method, the time-of-detection is on average just after the critical transition (no lead-time), with a mean time-of-detection of 82.2 for the Ext simulations (blue distribution Fig. 9(a)). As we consider a 12 consecutive point strategy, the points above the threshold begin on average at 70.22, thus, if a less strict constraint is used, the time-of-detection will on average be before the critical transition. There are few false detections with the steady state data (8 out of 500, shown in the green distribution) and 107 simulations of NExt data returned a false detection (purple distribution). The false positive rate found with the 2-sigma method is much lower than the high number of false positives returned with the other EWS based methods: Kendall’s *τ p*-value approach (Fig. 9(b)) and logistic composite (Fig. 9(c)). Although, the logistic method has a fairly predictive performance when tested on the Fix dataset (*FPR*_1_ = 0.214), it is worse when tested with the NExt data (*FPR*_2_ = 0.672, correctly classifying only 164 simulations). Even so, it does not perform as poorly as Kendall’s *τ p*-value method, which cannot distinguish the NExt simulations from the Ext simulations (Fig. 9(b)); this approach detected disease elimination in 496 (out of 500) NExt simulations and 497 (out of 500) Ext simulations.

**Fig 9.**
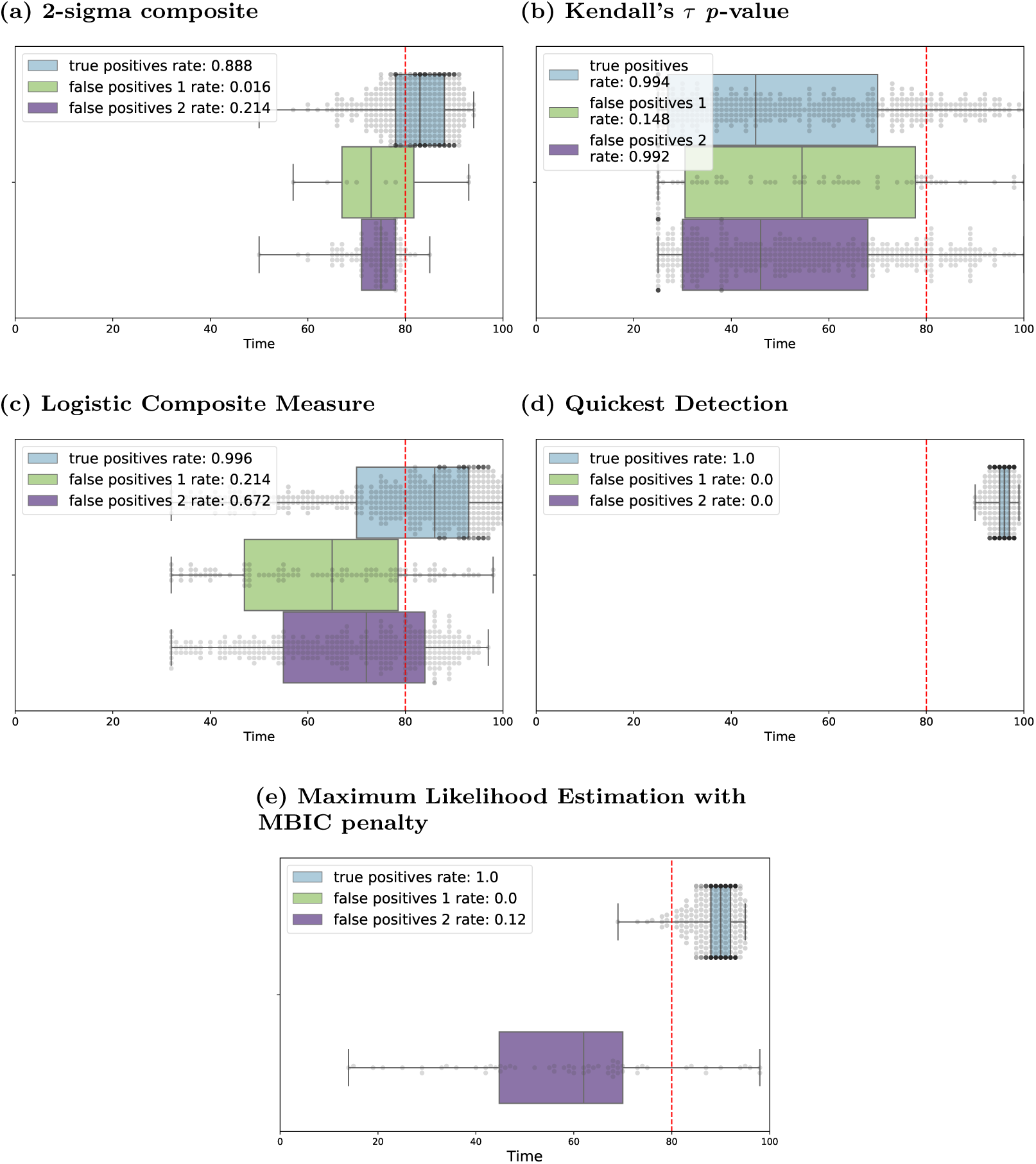
Time-of-detection from simulated data: for Ext data (blue boxplots), Fix data (green boxplots) and NExt data (purple boxplots). For each 500 simulation sets, a detection is observed following the threshold criteria of each method: (a) 2-sigma composite with CV and 12 consecutive points; (b) Kendall’s *τ p*-value with CV and 5 consecutive points; (c) logistic composite with InD and 21 consecutive points; (d) quickest detection with weights obtained from the MLE method, *A* = *log*(100) and 6 consecutive points and (e) MLE with the MBIC penalty criteria. Vertical red dashed line shows the time of the true bifurcation calculated for disease elimination. A list of EWSs and their acronyms used in this figure can be found in Supplementary Methods S2

The shape of the distribution of detection times is wider for Kendall’s *τ p*-value and the logistic composite compared to the 2-sigma method; where a narrower distribution suggests a higher confidence in the returned time-of-detection. However, the logistic composite method has a higher sensitivity (blue distribution Fig. 9(c)) and the mean time-of-detection is before the critical transition (mean 79 with a lead-time of 1 timestep). This implies that the Ext simulations started to cross the threshold on average at *t* = 58 (lead-time of 22 timesteps). This slight lead-time (with constraint) agrees with our findings in Fig. S2(a) where the power metric reduces for InD when only considering the time-series up to the bifurcation. However, for other indicators such as the combination of standard deviation, skewness and CV, the power metric did not decrease and therefore will have a larger lead-time and may be preferred.

In contrast, the change-point analysis based methods return a perfect sensitivity (score of 1, Fig. 9(d,e)) and a near perfect specificity for both null datasets. The MLE method with the MBIC penalty is given in Fig. 9(e), we find that the MBIC penalty reduces the largest number of false detections compared to the other penalties considered, shown in Fig. S4. The higher the penalty criteria, the fewer the number of NExt simulations incorrectly classified. The quickest detection approach does not return any false detections, although the time-of-detection with Ext data is on average at *t* = 95.78. This suggests that the quickest detection method is detecting disease extinction rather than when *R*_0_ crosses through 1, agreeing with our analysis of the power metric up to the bifurcation (Fig. 7). Further, only 10 out of 500 Ext simulations gave a time-of-detection before the critical transition for the MLE approach. Although we use change-point analysis to detect when the variance changes (an EWS detection), both MLE and quickest detection signal after the bifurcation, indicating that change-point analysis approaches do not give an early warning of the critical transition.

## Table Notes

The table provides the TPR, TNR_1_ and TNR_2_ for the most predictive statistic and number of consecutive points of each detection method. The best performing online method is highlighted in violet, the second best in blue and the MLE (offline method) is highlighted in grey. A result in bold font has a classification rate ≥ 0.75.

For the quickest detection method, we define the distributions *f* and *g* to be Gaussian and use a mean of zero (since the data are detrended) with the values of *σ*_1_ and *σ*_2_ found using the MLE approach. We estimate *σ*_1_ = 34 ≥2.4 = *σ*_2_ and for this distribution the results of quickest detection are in Fig. 9. However, when implementing the quickest detection method without using MLE to find the parameters, Carpenter *et al*. [24] suggested using the first 50 time-points of the time-series. We consider the variance of the first 25 time-points for a time-series of length 100 and find the range of estimated *σ*_1_’s to be [20.5, 53.4] (from the 500 simulations of Ext). By taking *σ*_2_ to be a reduction of *σ*_1_, we show in Fig. S5 how the quickest detection method performs for a different distribution. In particular, we find that for *σ*_1_ = 40 and *σ*_2_ = 10, the time-of-detection is earlier (nearer to the bifurcation), although the number of false positives increases.

In Table 1 we summarise the results of Fig. 9 for each time-of-detection method. We also present the sensitivity and specificity of each method when performed on time-series data of length: 20 (Table S1); 50 (Table S2) and 250 (Table S3). In particular, bold font in all tables indicates a robust score (*TPR, TNR*_1_ or TNR_2_) which is greater or equal to 0.75. The grey shading indicates that the MLE method is an offline approach, which should be accounted for in the comparison. Violet shading highlights the detection method which has the maximum sensitivity and specificity. In particular, we find that the quickest detection method performs best for all time-series lengths. The blue shading highlights any other suitable method. We find that the 2-sigma method is the only (non change-point analysis) approach that is robust for the detection of disease elimination. In particular, we find that the quickest detection method with different parameter values (Fig. S5) has a lower false positive rate than the EWS based methods for long time-series (lengths 100 and 250), however, for shorter time-series the 2-sigma method achieves the highest sensitivity and specificity. Thus, although the TPR and TNR_2_ are not robust (≥0.75) for the 2-sigma with time-series of length 20, it is the best performing method when the distributions of the quickest detection method are changed. However, we do not find the 2-sigma method to be a suitable approach for long time-series, as the TNR_2_ is near to 0.5 (false classification for 50% of the simulations).

**Table 1.**
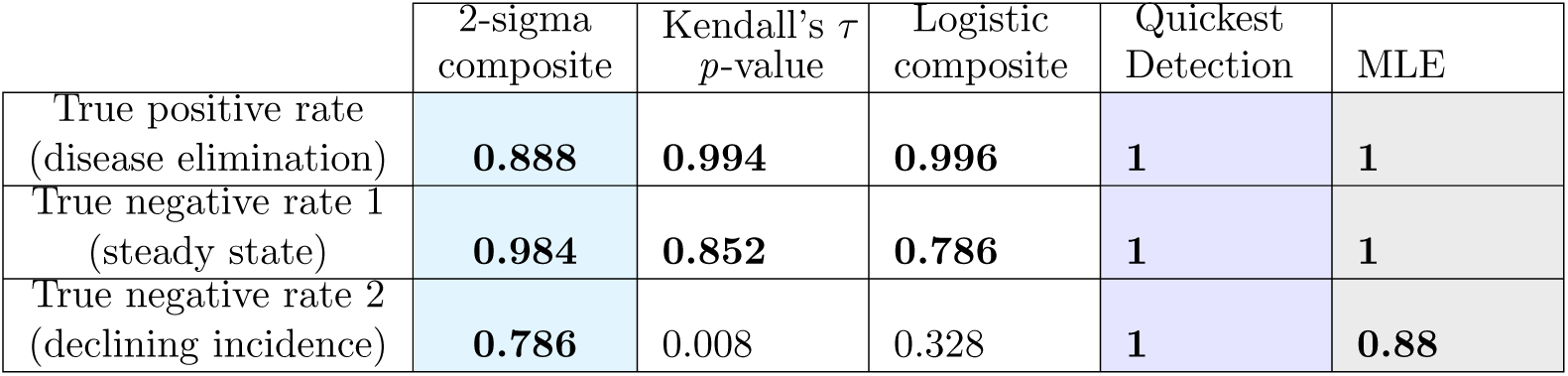
Time-series length 100. Statistic used (number consecutive points used): 2-sigma, CV (12); Kendall’s *τ*, CV (5); logistic, InD (21); quickest detection, *A* = *log*(100) (13).

## Discussion

A challenge of using EWSs to anticipate critical transitions is determining how to infer when an EWS becomes significant and how to measure how early a critical transition can be detected before it is reached. In this paper, we reviewed and validated the use of online detection algorithms from the EWS literature and assessed whether they can be applied to an infectious disease system for the detection of disease elimination.

Out of the three EWS based methods which we analysed, two of the methods (2-sigma threshold and changing *p*-value), had not been previously tested with a controlled environment. Communicating the performance of each algorithm for a controlled environment is clearly important for minimising the number of false detections when applying the algorithm in practice. In particular, many of the EWS based methods demonstrated that disease elimination could be distinguished from a disease at the endemic steady state. However, we found that if an algorithm is only tested on data described by a controlled environment at the steady state it may give a false impression on the reliability of the method. Ideally, online detection methods should not detect a signal in time-series data that have reduced reporting, which can occur if mass screening events stop or if a disease is no longer notifiable, given that this type of data may not be bifurcating. Here, we showed that it is also possible to distinguish disease elimination from a system of declining incidence which does not go through a critical transition. We found that the 2-sigma method was the only method which provided a robust and timely classification with this null dataset, although performance depended on which EWS was used. In particular, the specificity of all detection methods were markedly worse when using a more realistic null dataset. We consistently found that change-point analysis based methods reduced the false positive rate the most for both null datasets. However, we found that change-point analysis based approaches did not provide a lead-time of the critical transition. This work supports the use of change-point analysis to identify when a bifurcation occurred, such as with historical data, although we suggest that they are inappropriate for the detection of disease emergence, whereby a large lead-time is necessary.

Since epidemiological datasets are often of low temporal resolution, we considered how the methods performed for shorter time-series datasets. All online detection methods worsened when considering shorter time-series, we found that the specificity of all algorithms worsened when less than 50 data points were used, while the sensitivity could be maintained. Even so, the change-point analysis based methods were still robust in data poor settings. However, we found that if different input distributions were used with the quickest detection approach, the method was no longer suitable and was outperformed by the 2-sigma method for a time-series of length 20. Thus, a limitation of the quickest detection method is determining suitable distributions which describe the detrended data.

We found that the most reliable detections occurred when a consecutive point strategy was included. We demonstrated that by requiring multiple time-series points to be observed above the detection threshold, the performance of all algorithms greatly improved. The majority of false detections with the steady state data were sparse isolated points which crossed the threshold. For the more realistic null dataset, small clusters of points were observed above the threshold. Hence, using a consecutive point strategy would reduce the false positive rate. Here, we examined the number of consecutive points which would be optimal to minimise the classification errors. We noted that by including an additional constraint, the lead-time reduced, as multiple points were required before a detection was alarmed. We found that for some detection methods, a strategy using over 40% of the length of the time-series would be recommended as the best number of consecutive points, thus delaying the first possible time-of-detection by over 40%. Therefore, there is a trade off between whether a high specificity is necessary or a longer lead-time.

The advantage of using a combination of multiple EWSs has previously been discussed and the 2-sigma method and logistic composite method used a combination of EWSs. With the 2-sigma method we found that the most reliable indicator was a single indicator (CV), although the EWSs which provided the best lead-time were the combination CV + Va. Similarly, for the logistic method, the most reliable indicator was InD, however again a combination of EWSs provided an earlier detection. Additionally, InD performed best with a large number of consecutive points (around 20%) while a combination of EWSs worked well with fewer consecutive points (around 5%). A potential improvement for the 2-sigma method would be to include a weighted composition, rather than using an indicator function. We considered using the weights obtained in the logistic regression with the 2-sigma composite method, testing 2.244CV - 2.3Va (instead of CV-Va) and our initial findings showed that the TPR improved from 0.454 to 1 and the TNR_1_ from 0.872 to 1 (Fig. 10). However, the TNR_2_ decreased significantly from 0.87 to 0.084. Thus, further exploration is needed to find suitable weights of EWSs, such that the false positive rate with NExt data is minimised. One potential approach could be to train the logistic classifier using the NExt null dataset rather than the Fix dataset.

**Fig 10.**
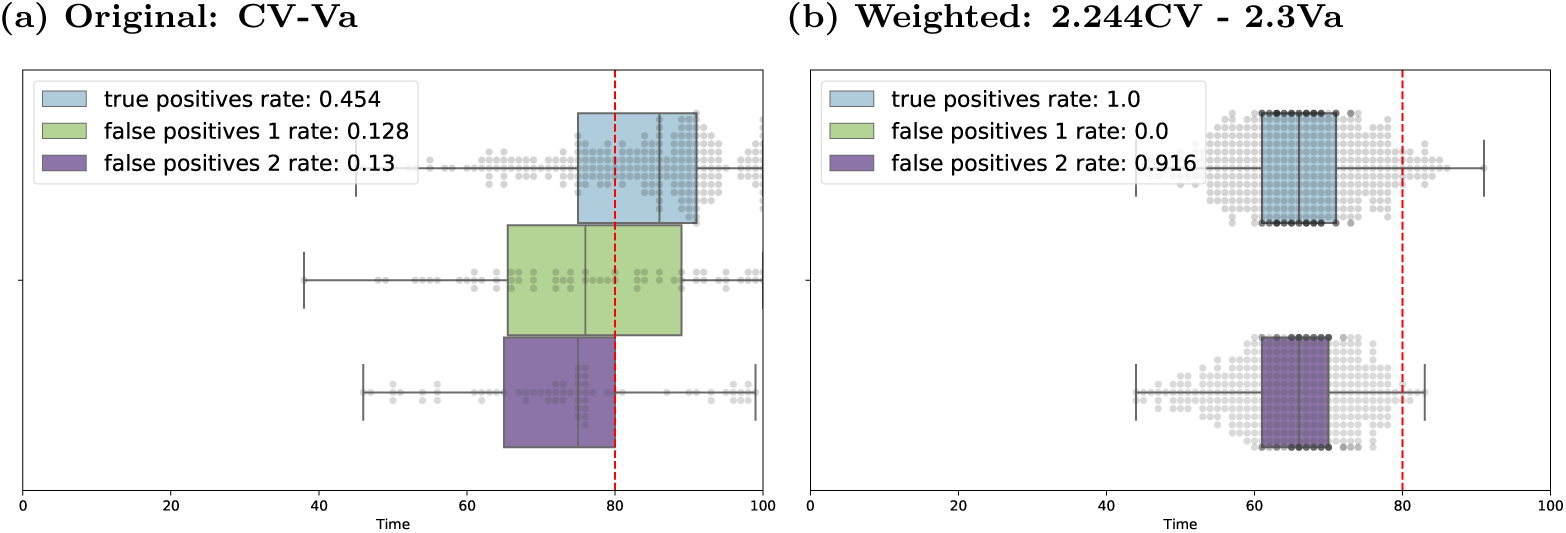
Weighted composite with the 2-sigma approach: For a time-series of length 100, the time-of-detection is calculated for the 500 simulations sets. We compare the composite CV-Va using the 2-sigma method and an 6 consecutive point constraint, with a weighted composite of the two indicators CV and Va. The weights are determined using the logistic regression from the logistic composite method, and the composition 2.244CV - 2.3Va is presented.

A limitation of the methods investigated here is whether they are truly model-independent. In particular, for the 2-sigma, Kendall’s *τ p*-value, quickest detection and MLE methods we used knowledge of the analytical behaviour of variance. We used a negative coefficient, right-tailed *p*-value and a decreasing *σ*_1_ ≥ *σ*_2_ constraint, respectively, to inform us that the variance of incidence should decrease on the approach to a critical transition. Even though the logistic composite method required a computationally exhaustive pre-processing element to train the algorithm, it did also obtain a negative coefficient for the variance, agreeing that the EWS decreases. The aim of EWSs is to provide model-independent approaches for informing critical transitions. However, due to discrepancies between systems when implementing these methods, in practice it is necessary to understand the expected behaviour of EWSs first.

These results strongly support the use of a consecutive point strategy as suggested by Clements *et al*. [32, 37]. We found that the original two-consecutive point strategy is insufficient for reducing the false positive rate with a more realistic null dataset. We analysed the best number of consecutive points which would improve the classification of each detection method and compared the reliability and lead-time of each detection method. Our results highlight that the optimal number of consecutive points depends on the EWS considered, the time-series length and the detection method used. We demonstrated that the 2-sigma method is the most robust EWS based detection method and found its strong performance was maintained for different lengths of time-series data. While, for retrospective analyses, we found that the quickest detection method performed better, providing the highest sensitivity and specificity. Further work is needed to address the suitability of the consecutive strategy with the detection of disease emergence and whether any of the detection methods considered will provide a sufficient lead-time with a disease emergence system. Finally, considerations for the operational use of detection algorithms remains an open topic. In particular, integrating EWSs into a decision theory framework to determine how decision makers should interpret the detection of an upcoming disease transition and assign a cost to a false detection of disease elimination.

## Data Availability

Data and code to reproduce results are available at https://github.com/ersouthall/Time-of-detection.

https://github.com/ersouthall/Time-of-detection

## Supporting information

**Supplementary Methods S1 Simulation Study** This section provides further details on the methods used to generate the synthetic data and the parameter values used in the simulation study (Table S1).

**Supplementary Methods S2 A description of how EWSs are computed** This section provides further details on how EWSs are computed on the time-series data, including a list of EWSs used in this paper (and their acronyms) in Table S2.

**Fig. S1 Power Metric Analysis:** for different time-series lengths considered (lengths: 20, 50, 100 and 250). Each heatmap shows the total power metric (2*TPR* − *FPR*_1_ − *FPR*_2_ ∈ [−2, 2]) for the two null models considered, where TPR is calculated as the proportion of disease elimination simulations which are successfully detected; FPR_1_ is the proportion of steady state simulations which are incorrectly detected and FPR_2_ is the proportion of declining incidence (but not bifurcating) simulations which are incorrectly detected. Each group of subplots shows the results for each detection method considered: (a) 2-sigma composite framework, (b) Kendall’s *τ p*-value methodology, (c) the logistic composite methodology and (d) the quickest detection approach. Each figure demonstrates how the performance of the total power metric changes when stricter constraints on the number of consecutive points required to cross the threshold are applied.

**Fig. S2 Power metric for time-series data up to the bifurcation point**. For a time-series of length 100 and bifurcation occurring at *t*^*^ = 80, the total power metric (2*TPR* − *FPR*_1_ − *FPR*_2_), is calculated over the time-series data up to the bifurcation. Results are shown for each detection method considered: (a, Kendall’s *τ p*-value); (b, logistic composite).

**Fig. S3 Number of consecutive points:** For each detection method, the “best” number of consecutive points are found from the ROC curves by selecting the number of consecutive points which minimises the classification error. Each scatter plot shows the best number of consecutive points as a percentage of the time-series length for each null dataset: triangular markers show NExt results and circular markers show Fix results. Results are shown for time-series of length (a) 20, (b) 50, (c) 100 and (d) 250 and for each online detection method: 2-sigma (blue); Kendall’s *τ p*-value (orange); logistic (green) and quickest detection (red).

**Fig. S4 Maximum likelihood estimation with different penalties:** For a time-series of length 100, the time-of-detection of Ext data (blue boxplots); Fix data (green boxplot) and NExt data (purple boxplots). For each 500 simulation set, the time-of-detection is calculated using the MLE approach and the alternative hypothesis is accepted following a penalty criteria. Penalty criteria considered: (a) Hannan-Quinn, (b) AIC, (c) Wilk’s Theorem, (d) BIC and (e) MBIC. The TPR, FPR_1_ and FPR_2_ are given in the legend.

**Fig. S5 Quickest detection performance for different distributions** *f* **and** *g*. Find 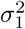 as the variance of the first 25 time-points of a time-series of length 100. For our 500 simulation sets, we find *σ*_1_ ∈ [20.5, 53.4]. In this example, we take *σ*_1_ = 40 and *σ*_2_ = 10 and use a 6 consecutive points constraint, as before.

**Table S1 Time-series length 20**. Statistic used (number consecutive points used): 2-sigma, CV (2); Kendall’s *τ*, CV (10); logistic, InD (3); quickest detection, *A* = *log*(20) (3).

**Table S2 Time-series length 50**. Statistic used (number consecutive points used): 2-sigma, CV (7); Kendall’s *τ*, CV (25); logistic, InD (10); quickest detection, *A* = *log*(50) (7).

**Table S3 Time-series length 250**. Statistic used (number consecutive points used): 2-sigma, CV (15); Kendall’s *τ*, CV (74); logistic, InD (49); quickest detection, *A* = *log*(250) (33).

## Acknowledgments

This work has been funded by the Engineering and Physical Sciences Research Council through the MathSys CDT [grant number EP/S022244/1]. The funders had no role in study design, data collection and analysis, decision to publish, or preparation of the manuscript

## Notes

### Competing Interest Statement

The authors have declared no competing interest.

