## Supplementary Information for "How early can an upcoming critical transition be detected?"

### 1 Supplementary Methods

This section provides further details on the methods used to generate the synthetic data, and compute the early warning signals of the time-series data. The code used to produce the data and generate the EWSs is available at `https://github.com/ersouthall/Time-of-detection`.

#### 1.1 Simulation Study

We generate stochastic simulations using the Gillespie algorithm (Gillespie, 1977) for an underlying Susceptible-Infectious-Susceptible (SIS) model. In this model, new infections are generated at a rate $\beta(t)SI/N$ and individuals recover from infection at a rate $\gamma I$. We study EWSs on incidence (new cases) data produced by the SIS model, where the system is slowly moving away from the endemic steady state and towards the disease free state. We use the version of the model proposed by (O'Regan and Drake, 2013) where the transmission rate

$$\beta(t) = \beta_0(1 - pt) \tag{1}$$

is gradually reducing through time, until the disease is unsustainable. Parameter values used for the simulations are given in Table 1, and the simulated data were initially aggregated into timepoints $\Delta t = 1$ years apart. We investigate the quantity of time-series data needed to provide robust EWSs, by increasing $\Delta t$ to reflect less frequent sampling of incidence data. We consider $\Delta t \in [1, 2, 5, 10, 25]$.

| Parameter | Value | |
|---|---|---|
| Initial transmission rate | $\beta_0$ | 1 |
| Recovery rate | $\gamma$ | 0.2 |
| Change in transmission | $p$ | 1/500 |
| Population size | $N$ | 10,000 |
| Initial number of infections | $I(0)$ | 0.8N |

Table 1: Simulation parameter values

We evaluate the detection methods with three different types of incidence data:

- Bifurcating data: **Ext** (extinct), the SIS model described above, with a decreasing transmission rate $\beta(t) = \beta_0(1 - pt)$ over time.

- Null data: **Fix** (fixed/endemic), the SIS model with a fixed value of $\beta(t) = \beta_0$, for all $t$.
- Null data: **NExt** (not extinct), as in Ext, but $\beta(t)$ stops decreasing at $t*$ when $\beta(t*) = 1.3\gamma$ and remains at $\beta(t) = 1.3\gamma$ for the rest of the simulation ($t > t*$).

#### 1.2 A description of how EWSs are computed

In practice, without the availability of replicates, statistics cannot be calculated over multiple realisations. Instead, a moving average approach is often used, whereby the statistic is calculated on a subset of the time-series of length $w$. The time evolution of the statistic is achieved by shifting the window, $w$, forward by one timepoint, so that each new subset contains the next timepoint and removes the first point of the previous subset. For example, the variance of a single time-series $\{x(t) : t \in [0, n-1]\}$, can be estimated using a right-edge window,

$$Var(x(t)) = \frac{1}{w-1} \sum_{t'=t-w}^{t} (x(t') - \hat{x}(t))^2,$$

$$\hat{x}(t) = \frac{1}{w} \sum_{t'=t-w}^{t} x(t'),$$

or a central-window,

$$Var(x(t)) = \frac{1}{w-1} \sum_{t'=t-\frac{w}{2}}^{t+\frac{w}{2}} (x(t') - \hat{x}(t))^2,$$

$$\hat{x}(t) = \frac{1}{w} \sum_{t'=t-\frac{w}{2}}^{t+\frac{w}{2}} x(t').$$

A right-edge window is often preferred as it does not use future data points in the estimation of the statistic at time $t$. In this paper, we use right-edge windows in our analysis and the numerical formulas for the statistical moments used in this paper are shown in Table 2.

| Statistic | Mathematical Definition | Numerical (N replicates) | Numerical (moving average) |
|---|---|---|---|
| Mean | $\mu_t = \mathbb{E}[X_t]$ | $\mu_R(t) = \frac{1}{N}\sum_{r=1}^{N} x_r(t)$ | $\mu_w(t) = \frac{1}{w}\sum_{t'=t-w}^{t} x(t')$ |
| Residuals | $y_t = X_t - \mu_t$ | $y_r(t) = x_r(t) - \mu_R(t)$ | $y_w(t') = x(t') - \mu_w(t), t' \in [t-w, t]$ |
| Variance (Va) | $\sigma_t^2 = \mathbb{E}[X_t^2] - \mathbb{E}[X_t]^2$ | $\sigma_R^2(t) = \frac{1}{N}\sum_{r=1}^{N} y_r(t)^2$ | $\sigma_w^2(t) = \frac{1}{w-1}\sum_{t'=t-w}^{t} y_w(t')^2$ |
| Coefficient of variation (CoV) | $\sigma_t/\mu_t$ | $\sigma_R(t)/\mu_R(t)$ | $\sigma_w(t)/\mu_w(t)$ |
| Index of dispersion (InD) | $\sigma_t^2/\mu_t$ | $\sigma_R(t)^2/\mu_R(t)$ | $\sigma_w(t)^2/\mu_w(t)$ |
| Skewness (SK) | $\mathbb{E}[y_t^3]/\sigma_t^3$ | $\frac{1}{N\sigma_R(t)^3}\sum_{r=1}^{N} y_r(t)^3$ | $\frac{1}{w\sigma_w(t)^3}\sum_{t'=t-w}^{t} y_w(t')^3$ |
| Kurtosis (KT) | $\mathbb{E}[y_t^4]/\sigma_t^4$ | $\frac{1}{N\sigma_R(t)^4}\sum_{r=1}^{N} y_r(t)^4$ | $\frac{1}{w\sigma_w(t)^4}\sum_{t'=t-w}^{t} y_w(t')^4$ |
| Autocovariance (Acov) | $Acov_t(\tau) = \mathbb{E}[y_t y_{t+\tau}]$ | $Acov_{t_R}(\tau) = \frac{1}{N}\sum_{r=1}^{N} y_r(t)y_r(t+\tau)$ | $Acov_{t_w}(\tau) = \frac{1}{w}\sum_{t'=t-w}^{t} y_w(t')y_w(t'+\tau)$ |
| Autocorrelation (AC) | $AC_t(\tau) = Acov_t(\tau)/(\sigma_t\sigma_{t+\tau})$ | $AC_{t_R}(\tau) = Acov_{t_R}(\tau)/(\sigma_R(t)\sigma_R(t+\tau))$ | $AC_{t_w}(\tau) = Acov_{t_w}(\tau)/(\sigma_w(t)\sigma_w(t+\tau))$ |
| Decay Time (DT) | $-\tau/\ln(AC_{t_R}(\tau))$ | $-\tau/\ln(AC_t(\tau))$ | $-\tau/\ln(AC_{t_w}(\tau))$ |

Table 2: List of early warning signals used in this paper (and their acronyms). Note: for the calculation of statistics on a moving window, the sample form is taken (degree of freedom is one).

### 2 Supplementary Figures

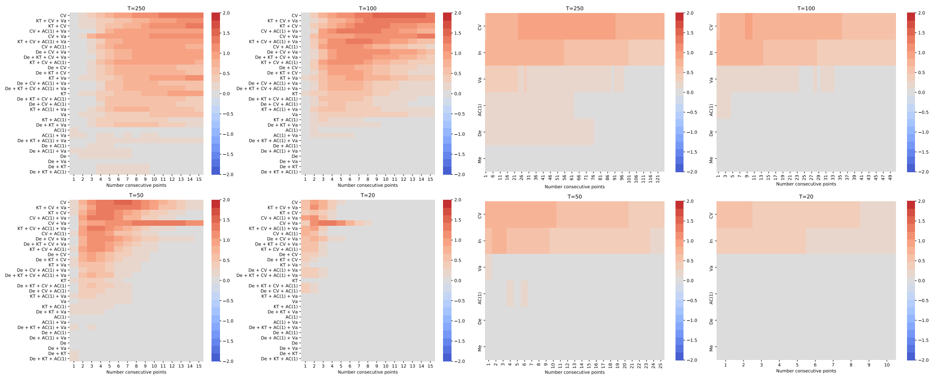

((a)) **2-sigma composite** ((b)) **Kendall's $\tau$ $p$-value**

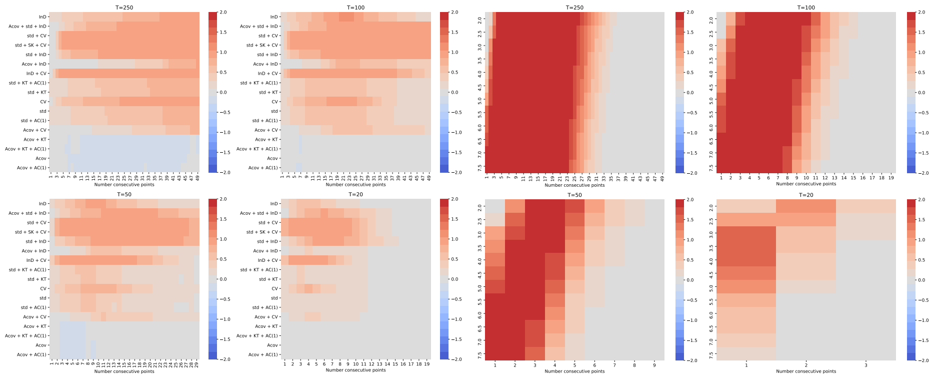

((c)) **Logistic composite** ((d)) **Quickest Detection**

Figure 1: **Power Metric Analysis:** for different time-series lengths considered (lengths: 20, 50, 100 and 250). Each heatmap shows the total power metric ($2TPR - FPR_1 - FPR_2 \in [-2, 2]$) for the two null models considered, where $TPR$ is calculated as the proportion of disease elimination simulations which are successfully detected; $FPR_1$ is the proportion of steady state simulations which are incorrectly detected and $FPR_2$ is the proportion of declining incidence (but not bifurcating) simulations which are incorrectly detected. Each group of subplots shows the results for each detection method considered: (a) 2-sigma composite framework, (b) Kendall's $\tau$ $p$-value methodology, (c) the logistic composite methodology and (d) the quickest detection approach. Each figure demonstrates how the performance of the total power metric changes when stricter constraints on the number of consecutive points required to cross the threshold are applied.

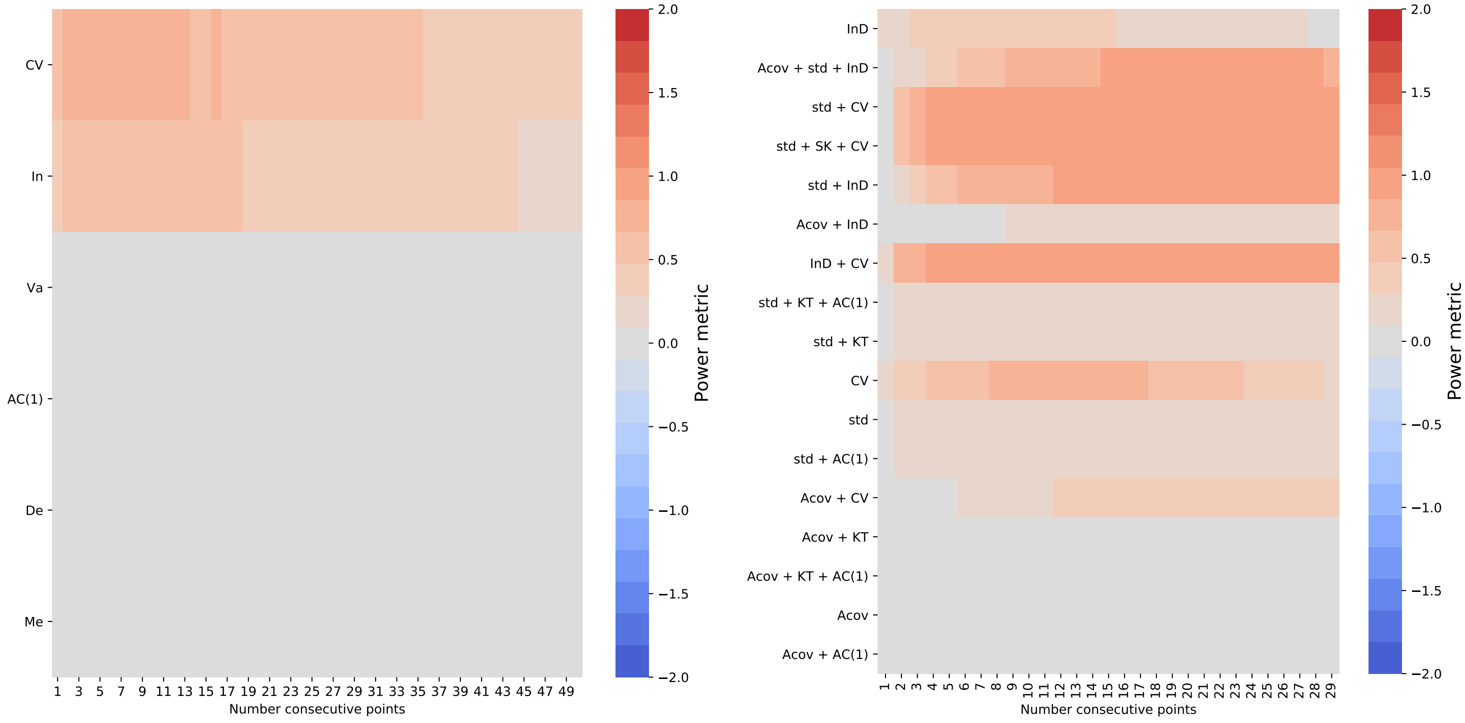


((a)) **Kendall's $\tau$ *p*-value** ((b)) **Logistic composite**

Figure 2: **Power metric for time-series data up to the bifurcation point.** For a time-series of length 100 and bifurcation occurring at $t^* = 80$, the total power metric $(2TPR - FPR_1 - FPR_2)$, is calculated over the time-series data up to the bifurcation. Results are shown for each detection method considered: (a, Kendall's $\tau$ $p$-value); (b, logistic composite).

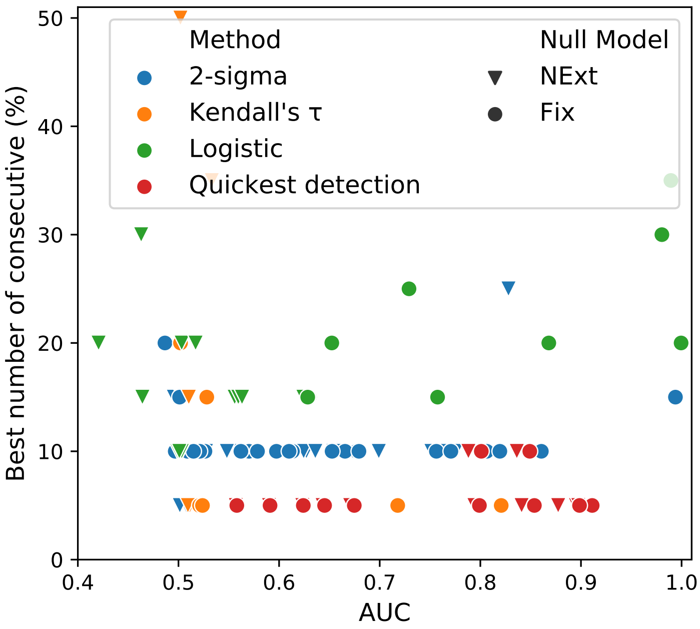


((a)) **Time-series length 20**

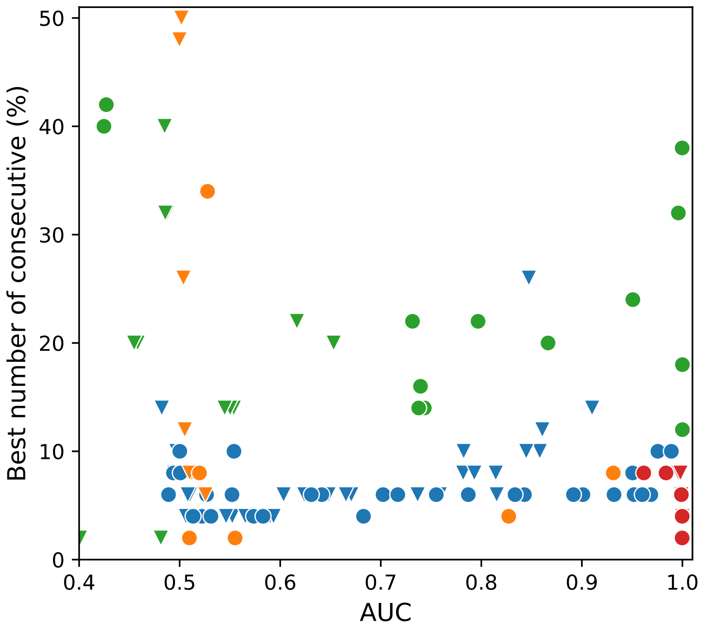


((b)) **Time-series length 50**

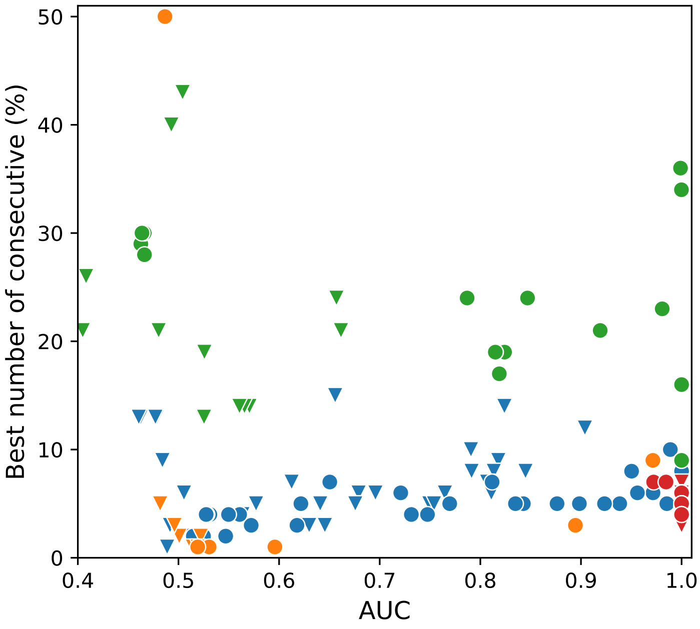


((c)) **Time-series length 100**

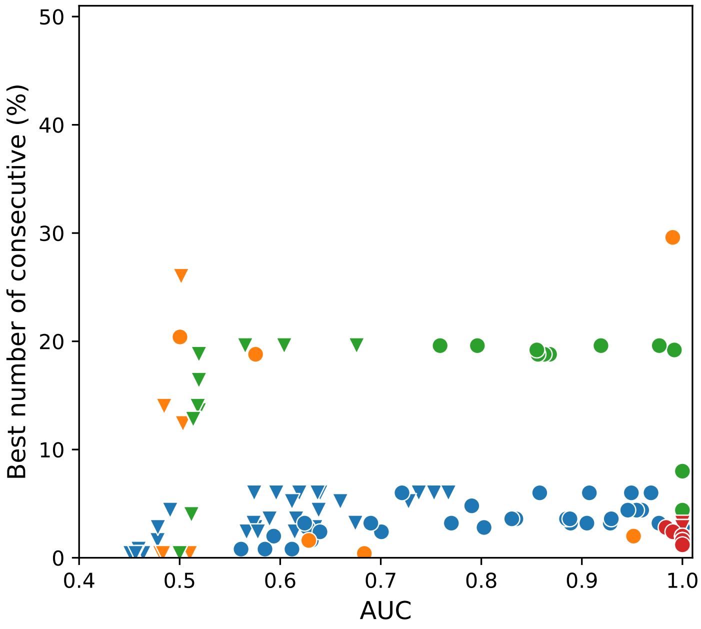


((d)) **Time-series length 250**

Figure 3: **Number of consecutive points:** For each detection method, the "best" number of consecutive points are found from the ROC curves by selecting the number of consecutive points which minimises the classification error. Each scatter plot shows the best number of consecutive points as a percentage of the time-series length for each null dataset: triangular markers show NExt results and circular markers show Fix results. Results are shown for time-series of length (a) 20, (b) 50, (c) 100 and (d) 250 and for each online detection method: 2-sigma (blue); Kendall's $\tau$ $p$-value (orange); logistic (green) and quickest detection (red).

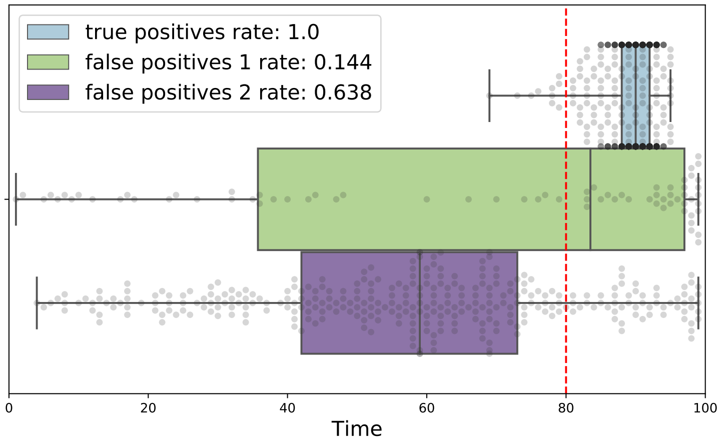


((a)) **Hannan-Quinn information criterion**

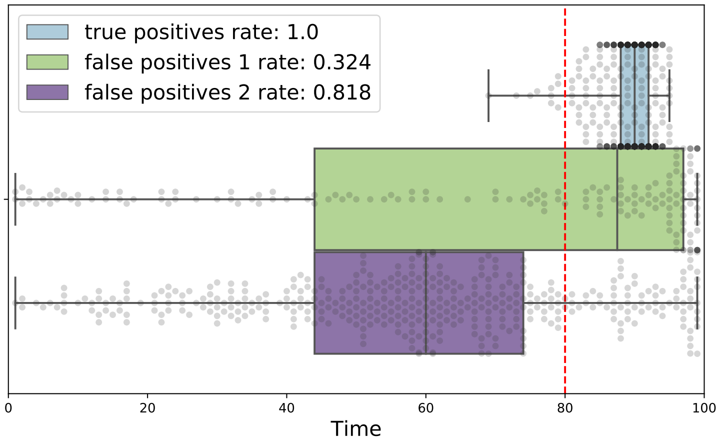


((b)) **Akaike information criterion**

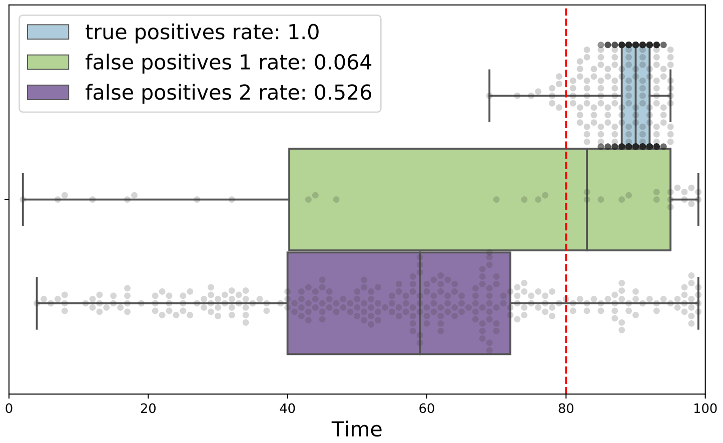


((c)) **Wilk's Theorem**

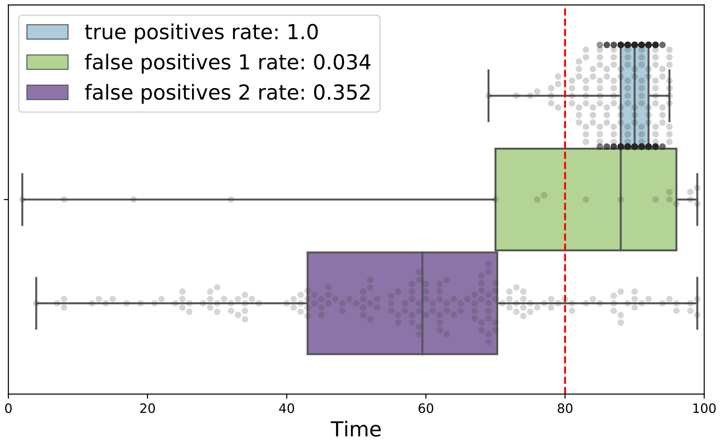


((d)) **Bayesian information criterion**

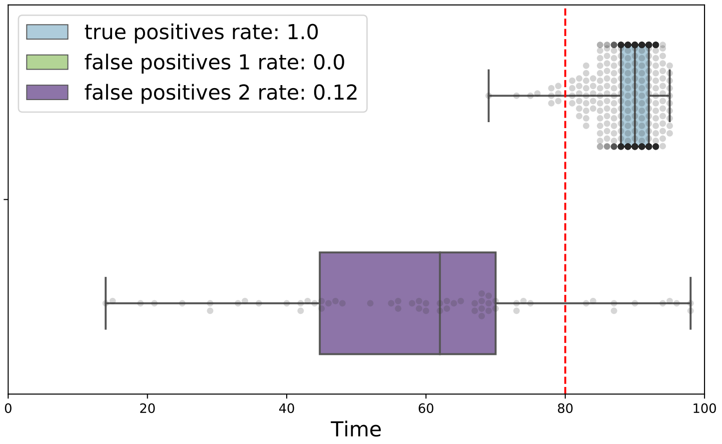


((e)) **Modified Bayesian information criterion**

Figure 4: **Maximum likelihood estimation with different penalties:** For a time-series of length 100, the time-of-detection of Ext data (blue boxplots); Fix data (green boxplot) and NExt data (purple boxplots). For each 500 simulation set, the time-of-detection is calculated using the MLE approach and the alternative hypothesis is accepted following a penalty criteria. Penalty criteria considered: (a) Hannan-Quinn, (b) AIC, (c) Wilk's Theorem, (d) BIC and (e) MBIC. The $TPR$, $FPR_1$ and $FPR_2$ are given in the legend.

| | 2-sigma composite | Kendall's $\tau$ $p$-value | Logistic composite | Quickest Detection | MLE |
|---|---|---|---|---|---|
| True positive rate (disease elimination) | 0.712 | 0.218 | **0.992** | **0.97** | **0.968** |
| True negative rate 1 (steady state) | **0.846** | **0.998** | 0.438 | **0.808** | **0.986** |
| True negative rate 2 (declining incidence) | 0.746 | **0.82** | 0.24 | **0.77** | **0.936** |

Table 3: **Time-series length 20**. Statistic used (number consecutive points used): 2-sigma, CV (2); Kendall's $\tau$, CV (10); logistic, InD (3); quickest detection, $A = log(20)$ (3).

| | 2-sigma composite | Kendall's $\tau$ $p$-value | Logistic composite | Quickest Detection | MLE |
|---|---|---|---|---|---|
| True positive rate (disease elimination) | **0.816** | 0.426 | **0.968** | **0.994** | **1** |
| True negative rate 1 (steady state) | **0.984** | **0.992** | 0.71 | **0.994** | **0.986** |
| True negative rate 2 (declining incidence) | **0.85** | 0.6 | 0.366 | **0.99** | **0.918** |

Table 4: **Time-series length 50**. Statistic used (number consecutive points used): 2-sigma, CV (7); Kendall's $\tau$, CV (25); logistic, InD (10); quickest detection, $A = log(50)$ (7).

| | 2-sigma composite | Kendall's $\tau$ $p$-value | Logistic composite | Quickest Detection | MLE |
|---|---|---|---|---|---|
| True positive rate (disease elimination) | **0.964** | 0.742 | **1** | **1** | **1** |
| True negative rate 1 (steady state) | **0.944** | **0.782** | **0.844** | **1** | **0.996** |
| True negative rate 2 (declining incidence) | 0.53 | 0.288 | 0.208 | **1** | 0.706 |

Table 5: **Time-series length 250**. Statistic used (number consecutive points used): 2-sigma, CV (15); Kendall's $\tau$, CV (74); logistic, InD (49); quickest detection, $A = log(250)$ (33).

**Table Notes:** Each table provides the $TPR$, $TNR_1$ and $TNR_2$ for the most predictive statistic and number of consecutive points of each detection method. This is demonstrated for different time lengths: 20 (Table 3), 50 (Table 4), 100 (Table **??**) and 250 (Table 5). The best performing online method is highlighted in violet, the second best in blue and the MLE (offline method) is highlighted in grey. A result in bold font has a classification rate $\geq 0.75$.

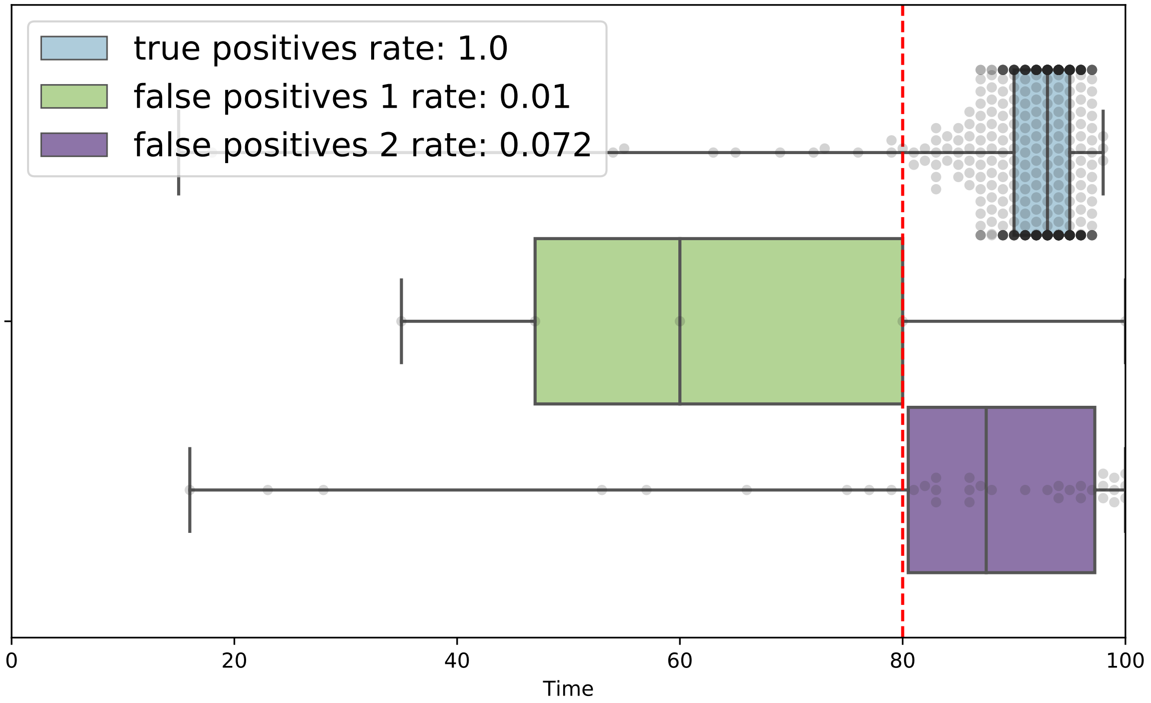


Figure 5: **Quickest detection performance for different distributions $f$ and $g$.** Find $\sigma_1^2$ as the variance of the first 25 time-points of a time-series of length 100. For our 500 simulation sets, we find $\sigma_1 \in [20.5, 53.4]$. In this example, we take $\sigma_1 = 40$ and $\sigma_2 = 10$ and use a 6 consecutive points constraint, as before.
